# Predicting the Tumor Microenvironment Composition and Immunotherapy Response in Non-Small Cell Lung Cancer from Digital Histopathology Images

**DOI:** 10.1101/2024.06.11.24308696

**Authors:** Sushant Patkar, Alex Chen, Alina Basnet, Amber Bixby, Rahul Rajendran, Rachel Chernet, Susan Faso, Prashant A. Kumar, Devashish Desai, Ola El-Zammar, Christopher Curtiss, Saverio J. Carello, Michel Nasr, Peter Choyke, Stephanie Harmon, Baris Turkbey, Tamara Jamaspishvili

## Abstract

Immune checkpoint inhibitors (ICI) have become integral to treatment of non-small cell lung cancer (NSCLC). However, reliable biomarkers predictive of immunotherapy efficacy are limited. Here, we introduce HistoTME, a novel weakly supervised deep learning approach to infer the tumor microenvironment (TME) composition directly from histopathology images of NSCLC patients. We show that HistoTME accurately predicts the expression of 30 distinct cell type-specific molecular signatures directly from whole slide images, achieving an average Pearson correlation of 0.5 with the ground truth on independent tumor cohorts. Furthermore, we find that HistoTME-predicted microenvironment signatures and their underlying interactions improve prognostication of lung cancer patients receiving immunotherapy, achieving an AUROC of 0.75[95% CI: 0.61-0.88] for predicting treatment responses following first-line ICI treatment, utilizing an external clinical cohort of 652 patients. Collectively, HistoTME presents an effective approach for interrogating the TME and predicting ICI response, complementing PD-L1 expression, and bringing us closer to personalized immuno-oncology.

## Introduction

Lung cancer is the leading cause of cancer-related mortality globally, of which non-small cell lung cancer (NSCLC) is the most common histological subtype^1^. In recent years, immune checkpoint inhibitors (ICI) have radically transformed the prognosis of clinically advanced NSCLC. Biomarkers such as Programmed Death Ligand 1 (PD-L1) expression have been clinically approved to identify patients who may respond to ICI. Yet, even after selecting patients based on PD-L1 expression, response rates to treatment can vary widely, ranging from 17-49% in patients with Tumor Proportion Score (TPS) > 1%^2^. This variability stems from the lack of standardized testing criteria for PD-L1, limiting its robustness as a predictive biomarker^3^. The clinical utility of other biomarkers such as the Tumor Mutational Burden (TMB) has been explored in several different clinical trials^4–7^. However, both PDL-1 expression and TMB fail to encapsulate various tumor microenvironmental features influencing ICI responses^8–10^. Hence, there is a clinically unmet need for additional predictive biomarkers capturing both tumor and microenvironmental factors associated with ICI responses.

In recent years, several new multiplex tissue imaging and spatial transcriptomics technologies have been used for profiling the tumor microenvironment (TME) of patients in unprecedented detail^11–19^. However, they are quite expensive, which does not allow them to be implemented for wider use in a clinical setting. Hematoxylin and Eosin (H&E)-stained pathology slides, on the other hand, are relatively cheap and easily accessible in any pathology labs. These slides hold a wealth of TME-related information that can be unlocked with the help of Artificial Intelligence (AI)^20^. One of the first large-scale attempts to characterize the TME of patients from H&E slides was the work of Saltz etal, who mapped the abundance and spatial distribution of tumor infiltrating lymphocytes (TILs) across 23 different cancer types^21,22^. Graham et al developed Hover-Net^23^, a pan-cancer nuclei segmentation and classification neural network that enables single-cell quantification of tumor, stroma and lymphocyte populations from H&E slides. More recently, Diao et al developed a collection of supervised machine learning (ML) methods to quantify 607 human interpretable TME features from histopathology images^24^. While these approaches are extremely valuable, they are limited by availability of relevant pixel-level annotations from expert pathologists, which are time and resource consuming to generate. To overcome these limitations, several research groups have alternatively proposed the use of weakly-supervised deep learning models, which can be trained to perform various downstream computational pathology tasks such as tumor subtyping and prognosis^25–30^ without any region or pixel-level annotations.

Building on these recent AI-based advances, we introduce HistoTME: a weakly supervised multi-task learning approach to infer the TME composition of patients from routinely collected pathology slides. Unlike previous approaches, HistoTME harnesses recently developed digital pathology foundation models to infer the expression of specific TME signatures, capturing the TME composition, without relying on any pathologist annotations or additional immunohistochemistry (IHC) data as labels. HistoTME is trained in a weakly supervised fashion utilizing matched whole slide H&E and bulk transcriptomics data of 865 NSCLC patients from The Cancer Genome Atlas (TCGA), validated on matched whole slide H&E and bulk transcriptomic data of 333 NSCLC patients from the Clinical Proteomic Tumor Analysis Consortium (CPTAC) and tested on whole slide H&E and IHC data from 82 NSCLC patients that had complete surgical resection at SUNY Upstate Medical University. We further demonstrate the clinical utility of HistoTME predictions by retrospectively analyzing needle biopsy specimens and clinical outcome data from an additional 570 NSCLC patients from the SUNY Upstate cohort treated with either chemotherapy or immune checkpoint inhibitors. Importantly, we show that HistoTME AI scores complement low PD-L1 expression and can identify more patients responding to immune checkpoint inhibitor therapy. Taken together, HistoTME presents a versatile and accessible tool for unravelling the complex dynamics of TME, leading to improved risk stratification and management of NSCLC patients.

## Results

### Overview of HistoTME

HistoTME is a deep learning model trained to predict the average normalized gene expression levels of 30 cell type-specific TME signatures from whole slide H&E images, collectively providing a comprehensive profile of the TME composition^31,32^ **(Figure 1A).** HistoTME was trained using whole slide images (WSI) of H&E staining and matched bulk transcriptomics data from the TCGA-NSCLC cohort (N=865 patients) and validated it using an external cohort of 333 NSCLC patients from CPTAC using the same data modalities **(Supplementary Figure 1)**. The HistoTME model consists of two main components: a frozen feature extraction component and a trainable attention-based multiple instance learning (AB-MIL) component^33^. In our efforts to efficiently train HistoTME, we explored three state-of-the-art open-source foundational models— CTransPath, RetCCL, and UNI^25,26,30^—as potential feature extractors. Additionally, we conducted experiments with two distinct approaches for AB-MIL: a single-task approach featuring a unique attention and multilayer perceptron (MLP) head for each TME signature, and a multitask approach, which incorporates a shared attention head for functionally related TME signatures but maintains separate MLP heads for each individual TME signature **(Fig. 1B)**.

**Figure 1:**
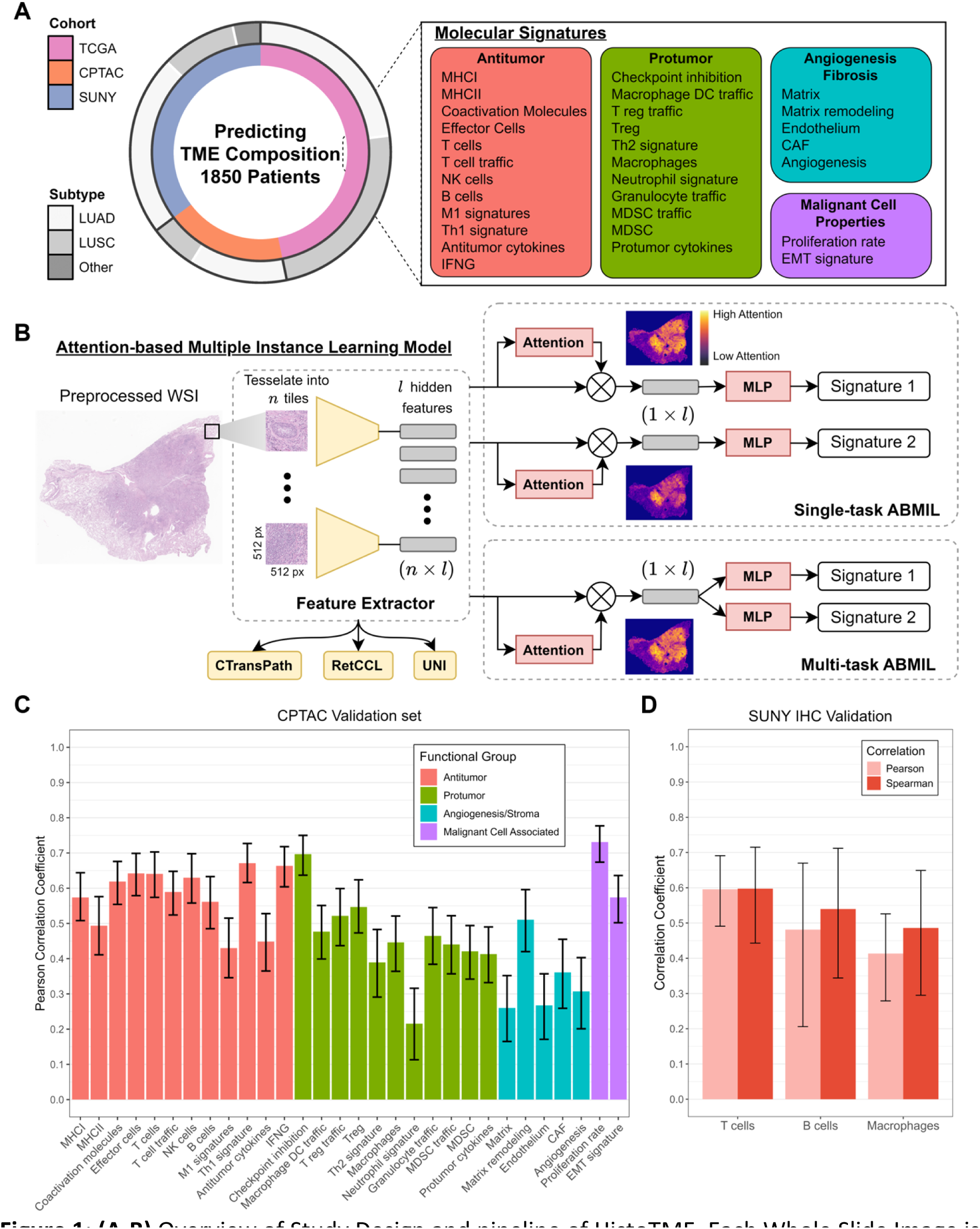
**(A-B)** Overview of Study Design and pipeline of HistoTME. Each Whole Slide Image is tessellated into smaller tiles and preprocessed by a pretrained digital pathology foundation model to extract meaningful tile embeddings. The tile embeddings generated by the foundation model are then provided as input to an attention-based multiple instance learning (AB-MIL) module followed by a multi-layer perceptron head (MLP), which learns to predict expression levels to 30 tumor microenvironment-related molecular signatures. Overall, to develop HistoTME we experiment with three open-source foundation models - CTransPath, RetCCL, and UNI^21,22,26^ - and two configurations of AB-MIL: single task AB-MIL, where the predictions of each signature are optimized separately, and multi-task AB-MIL, where predictions of functionally related signatures are jointly optimized. The signature prediction performance of each foundation model coupled with each configuration of AB-MIL is shown on held out CPTAC validation data in Supplementary Figure 3. Overall, the UNI foundation model + multitask AB-MIL produces the most accurate predictions and is hence chosen as the final version of HistoTME **(C)** Pearson correlations between the ground truth expression levels of each patient derived from bulk transcriptomics and predicted expression levels of each patient derived from the final version of HistoTME (UNI+multi-task AB-MIL) on the held out CPTAC validation cohort. **(D)** Pearson and Spearman correlations between the cell type abundance of each patient, defined as the number of marker positive cells per mm^2^ from immunohistochemistry (IHC) slides, and the predicted cell type-specific signature expression levels of each patient derived from final version of HistoTME (UNI+multitask AB-MIL) is shown on the external SUNY Upstate test cohort. Error bars represent the 95% confidence intervals. Cell type abundances were estimated from whole slide immunohistochemistry images using QuPath v0.5.0 cell detection and classification algorithms with default parameter settings. TME = tumor microenvironment; LUAD = lung adenocarcinoma; LUSC = lung squamous cell carcinoma; MLP = multilayer perceptron

### HistoTME accurately infers the TME composition of NSCLC patients from histopathology images

Of all versions of HistoTME that were explored, we observed that the version utilizing multi-task AB-MIL + the UNI foundation model produced the most accurate predictions when compared with the ground truth, achieving an average Pearson correlation coefficient of 0.50 **(Figure 1C**, **Supplementary Figure 2A)**. The performance of single-task AB-MIL + UNI was slightly worse than the performance of multi-task AB-MIL + UNI. However, both single-task AB-MIL + UNI and multi-task AB-MIL + UNI significantly outperformed other versions of HistoTME for predicting antitumor and protumor immune signatures while displaying similar performance for other signature prediction tasks **(Supplementary Figure 3)** Therefore, we settled on multitask AB-MIL+UNI as the final version of HistoTME. Having validated the accuracy of HistoTME on CPTAC-NSCLC data, we next tested HistoTME on whole slide H&E images of 82 NSCLC patients from SUNY Upstate Medical University, which had serial immunohistochemistry (IHC) performed on surgical resection specimens for immune cell panel: T cells (CD3, CD4, DC8), B cells (CD20) and Macrophage (CD163) markers using adjacent serial sections **(Supplementary Figure 1)**. Overall, we found that HistoTME predicted expression levels were correlated with the abundance of each cell type derived from IHC, achieving Pearson correlations of 0.60 [95% CI: 0.49-0.69] for T cells, 0.48 [95% CI: 0.21-0.67] for B cells, and 0.41 [95% CI: 0.28-0.53] for macrophages, and Spearman correlations of 0.60 [95% CI: 0.44-0.72] for T cells, 0.54 [95% CI: 0.34-0.71] for B cells, and 0.49 [95% CI: 0.30-0.65] for macrophages **(Figure 1D**, **Supplementary Figure 2B)**. Spearman correlations were included due to the uncertain linear correlation between gene expression and IHC-measured protein abundance^34^.

We then trained a simple unsupervised model on the TCGA + CPTAC cohorts to cluster NSCLC patients into distinct subgroups based on their predicted TME signatures **(Figure 2A**, **see Methods)**. This model effectively identifies two main clusters recapitulating the Immune-Inflamed and Immune-Desert phenotypes^35–37^**(Figure 2B**, **Supplementary Figure 4)**. Subsequently, we applied this two-cluster model to the entire institutional SUNY Upstate cohort, including patients with core needle biopsies (N=652 patients) **(Figure 2C)**. Upon examining the feature importance scores assigned to each TME signature by the trained two-class classification model **(Figure 2D)**, we found that the following signatures: T cell traffic, Antitumor Cytokines, MDSC(myeloid-derived suppressor cells), Co-activation molecules and Macrophage/Dendritic Cell Traffic were among the top 5 TME signatures driving the distinction between the Immune-Inflamed and Immune-Desert clusters. Interestingly, in line with these results, we noted substantial differences in the abundances of T cells, B cells and macrophages, when observed through IHC, among patients predicted to be either in the Immune Inflamed or in the Immune Desert cluster **(Figure 2E).**

**Figure 2.**
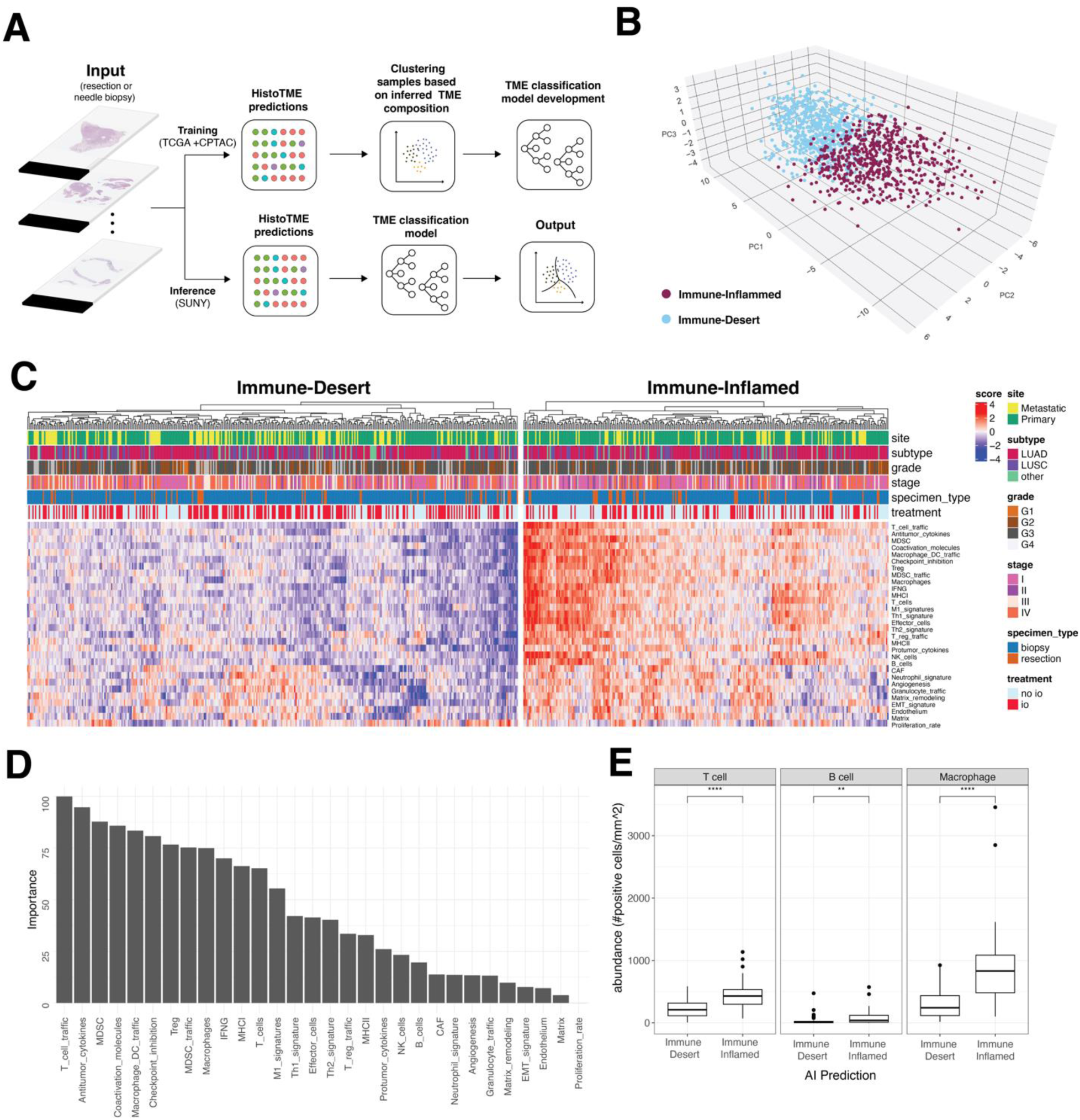
**(A)**: Overview of the computational pipeline to classify patients into distinct clusters based on their H&E-predicted TME composition. H&E stained digitized tumor samples from TCGA+CPTAC are processed by HistoTME and subsequently clustered into two clusters based on partition around medoid (PAM) clustering and a Random Forest classification model that is trained on cluster membership data **(B)** 3D PCA plot visualizing the two distinct clusters of TCGA + CPTAC NSCLC patients: Immune Inflamed and Immune Desert, based on their HistoTME-inferred TME profiles. **(C)** Heatmap depicting the H&E-predicted TME composition and clinical attributes of NSCLC patients from the SUNY cohort. Patients were classified into Immune Inflamed cluster or Immune Desert cluster using a two class classification model (Random Forest) trained on TCGA+CPTAC data. **(D)** Random forrest-derived feature importance rankings of TME signatures driving the distinction between the Immune Inflamed and Immune Desert cluster.**(E)** Immunohistochemistry-derived T cell, B cell and Macrophage abundances, defined as number of marker positive cells per mm^2^, in cases belonging to the immune-inflamed or immune desert cluster. Cell type abundances were quantified from whole slide immunohistochemistry images using QuPath v0.5.0 positive cell detection and quantification pipline. Statistical significance between groups was determined by non-parametric Wilox rank sum test (***: p-value < 0.001, ****: p-value < 0.0001)

To understand the histopathological features influencing HistoTME predictions, we generated attention maps for all 652 patients within the SUNY Upstate cohort. In general, HistoTME attends to different areas of the TME to estimate the expression of antitumor, pro-tumor, angiogenesis/stroma and malignant cell signatures, with the exception of antitumor and protumor immune signatures **(Supplementary Figure 5).** We additionally performed a qualitative review of four randomly chosen resection cases (2 Immune-Inflamed, 2 Immune-Desert) with the help of a board-certified pathologist **(Figure 3 and 4**, **Supplementary Figures 6 and 7)**. Overall, for immune-inflamed cases, which have relatively high predicted expression levels of antitumor and protumor immune signatures, HistoTME assigns great attention to regions abundant with lymphocytes and the formation of lymphocytic aggregates around tumor-stroma boundaries. For, immune desert cases, which have relatively low predicted expression levels of antitumor and protumor immune signatures, HistoTME assigns great attention to regions containing solid areas of more pleomorphic cells, and in addition, dense fibrotic areas within and around the tumor periphery. Collectively, these results suggest that HistoTME effectively captures the TME composition of tumors from H&E slides.

**Figure 3.**
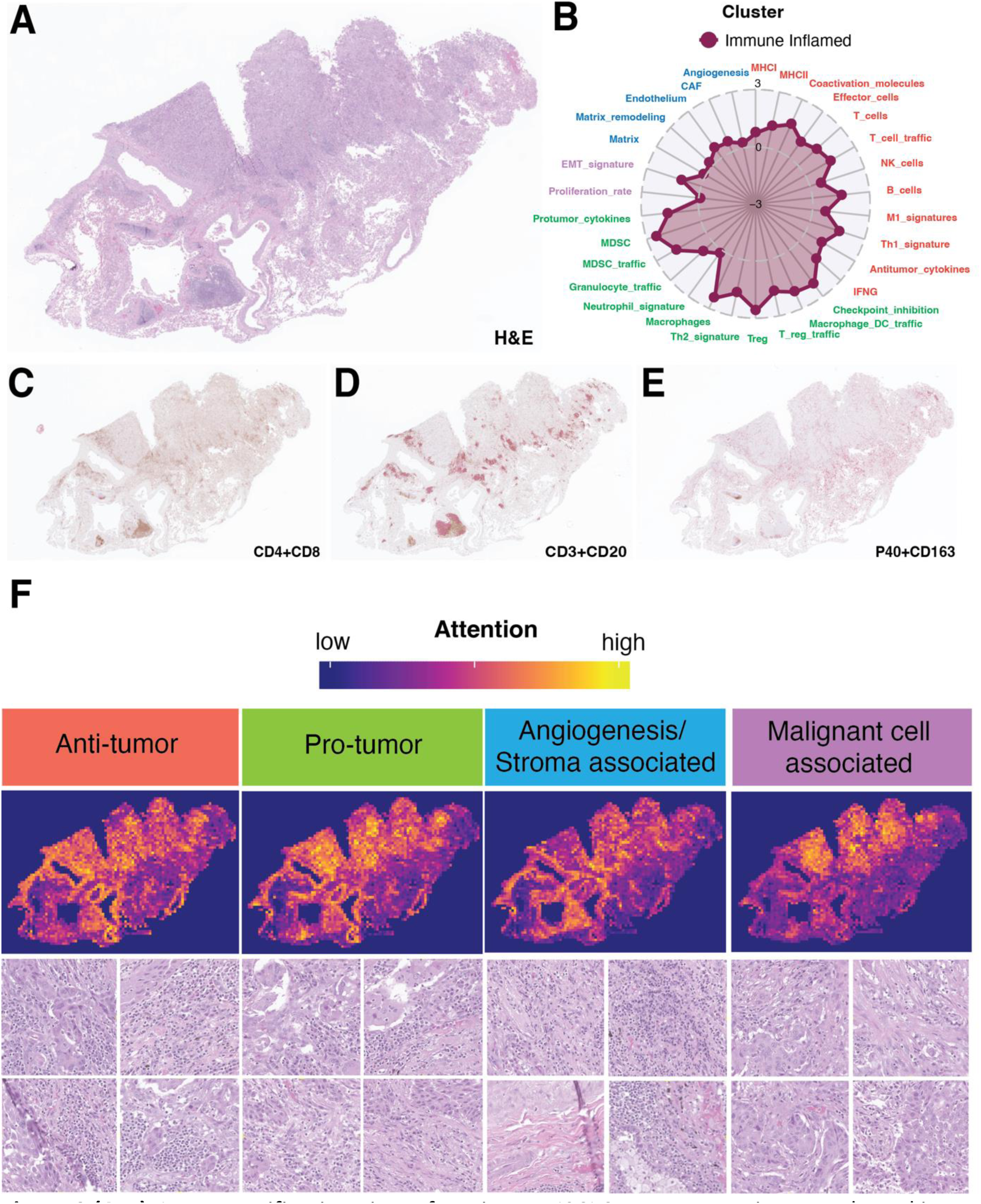
**(A-B)**: Low magnification view of a primary NSCLC tumor resection sample and its predicted TME signature profile. **(C-E)** Matched whole slide immunohistochemistry images of the same tumor sample dual stained for CD4 (brown)+CD8 (magenta), CD3 (brown)+CD20(magenta), and P40 (brown)+CD163 (magenta) markers respectively. (F) HistoTME generated attention maps for each attention head. Below each whole slide attention map are 4 high magnification image tiles (50×50 µm) randomly sampled from high attention areas. Supplementary Figure 6 shows another related example along with higher magnification image tiles randomly sampled from high attention areas.

**Figure 4.**
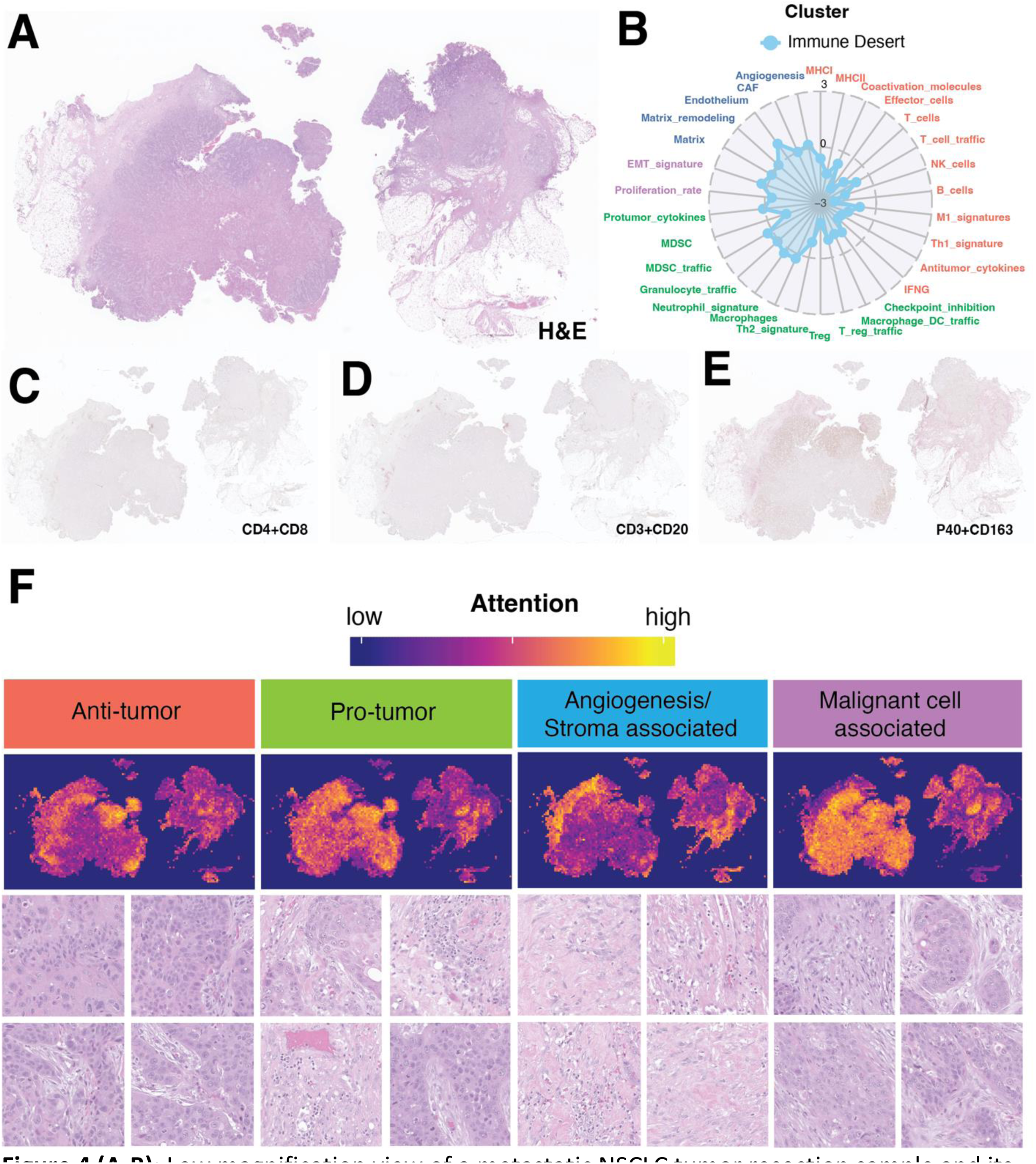
**(A-B)**: Low magnification view of a metastatic NSCLC tumor resection sample and its predicted TME signature profile. **(C-E)** Matched whole slide immunohistochemistry images of the same tumor sample dual stained for CD4 (brown)+CD8 (magenta)markers, CD3 (brown)+CD20(magenta) markers, and P40 (brown)+CD163 (magenta) markers respectively. **(F)** HistoTME generated attention maps for each attention head. Below each whole slide attention map are 4 representative high magnification image tiles (50×50 µm) sampled from high attention areas. Supplementary Figure 7 shows another related example along with higher magnification image tiles randomly sampled from high attention areas.

### Association between the tumor microenvironment status and survival outcomes of NSCLC patients treated with immune checkpoint inhibitors

We next conduct a retrospective analysis to assess how the TME status (Inflamed vs Desert), inferred from H&E slides, relates with survival outcomes of NSCLC patients from the SUNY Upstate cohort treated with immune checkpoint inhibitor therapy. All patients that underwent independent PD-L1 IHC testing with PD-L1 > 1% were considered for treatment with immune checkpoint inhibitors with a total of 292 patients ultimately receiving either an anti-PD1 or PD-L1 inhibitor as monotherapy or in combination with chemotherapy. Overall, 77% of these 292 patients had PD-L1 >= 1% and 50% had metastatic disease (stage IV). A detailed summary of the clinical cohort is available in **Supplementary Table 1**.

Interestingly, we found that the TME status is particularly predictive of overall survival of patients receiving ICIs as first line of therapy (**Figure 5A**, log-rank test p-value = 0.0012, HR = 0.53 [95% CI: 0.36-0.78]), especially when administered in combination with chemotherapy (First line ICI+chemo log-rank test p-value: 0.00067, HR = 0.39[95% CI: 0.22-0.68], First line ICI monotherapy log-rank test p-value: 0.22, HR = 0.68[95% CI: 0.37-1.26]; **Supplementary Figure 8**). Importantly, TME status remains predictive of overall survival of first line ICI-treated patients even after accounting for differences in clinical stage (HR = 0.52[95% CI: 0.35-0.77], p-value: 0.0012). The TME status, however, is less effective at predicting overall survival when considering all lines of ICI-treated patients (**Supplementary Figure 9A**, log-rank test p-value: 0.02, HR = 0.7 [95% CI: 0.52-0.95]). When looking at PD-L1 expression, patients with PDL1 expression >= 50% and receiving ICI treatment as first line of therapy showed markedly improved survival compared to those with PD-L1 1-49% or < 1% (**Figure 5B**, log rank test p-value = 0.0059, HR = 0.62 [95% CI: 0.37-1.04]). PDL1 expression was however not predictive of overall survival when considering all lines of ICI-treated patients (**Supplementary Figure 9B**, log rank test p-value = 0.13, HR = 0.9 [95% CI: 0.68 – 1.18]). When looking at progression-free survival (PFS), we observe that both the TME status and PD-L1 expression are primarily predictive of PFS at first line ICI therapy (TME status: log rank test p-value = 0.0037, HR = 0.59[95% CI: 0.41-0.85]; PD-L1 expression: log rank test p-value = 0.003, HR = 0.55[95% CI: 0.35-0.88]; **Supplementary Figure 10).** When performing additional subgroup analysis of patients receiving first line ICI treatment, we find that the H&E-inferred TME status is primarily predictive of overall survival outcomes of PD-L1 absent (<1%) patients (**Fig. 5C**; log rank test p-value = 0.08, HR = 0.4[95% CI: 0.13-1.18]), and PD-L1 low (1-49%) patients (**Fig. 5D**; log rank test p-value = 0.009, HR = 0.44[95% CI: 0.23-0.82]) but not PD-L1 high (>= 50%) patients (**Figure 5E**; log rank test p-value = 0.85, HR = 0.94[95% CI: 0.48-1.84]).

**Figure 5:**
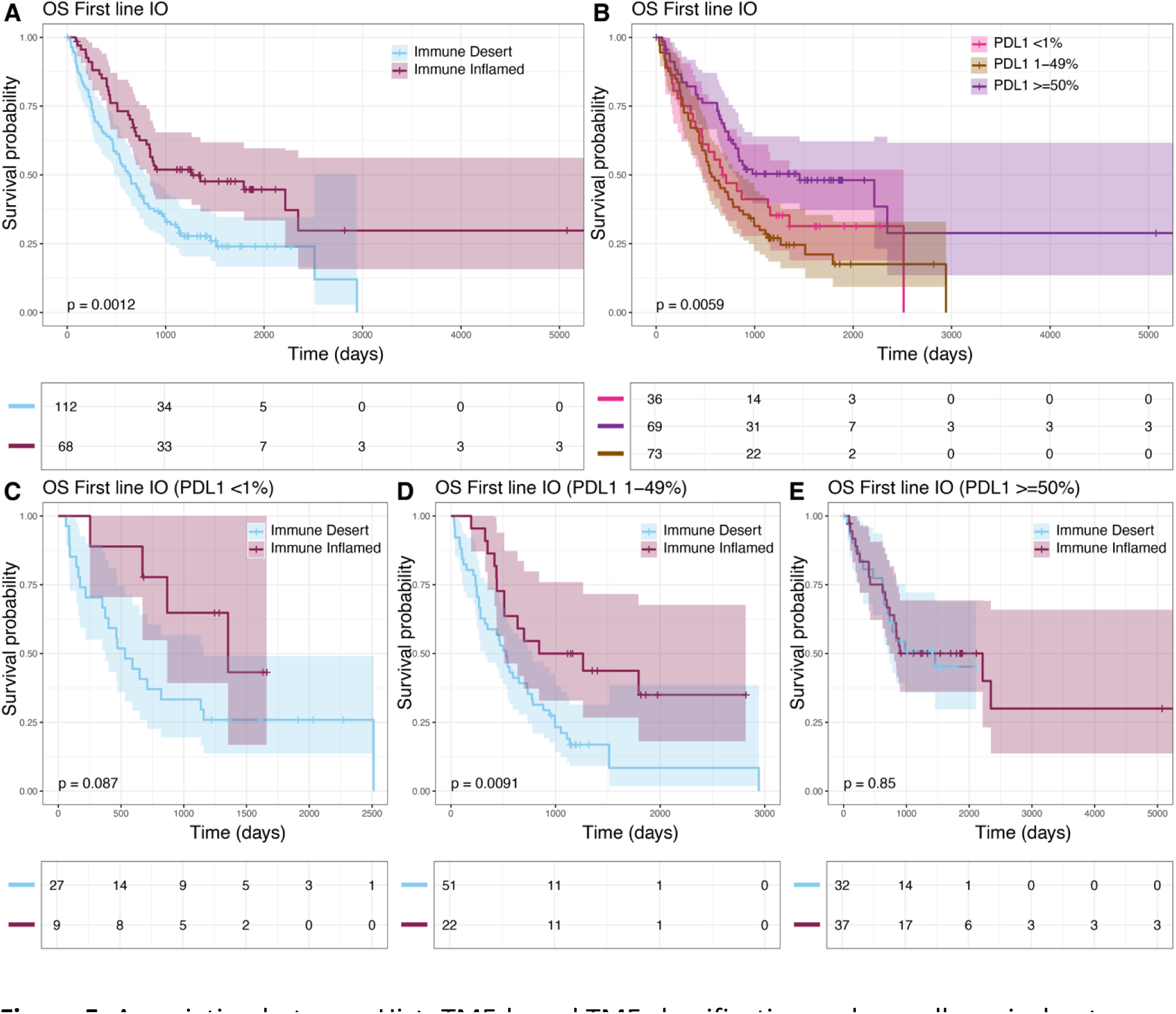
Association between HistoTME-based TME classification and overall survival outcomes of SUNY NSCLC patients treated with first-line anti-PD1/PD-L1 therapy (first-line IO patients). **(A)** Kaplan Meier plot depicting overall survival−defined as time from date of diagnosis to date of death −of patients that received first-line anti-PD1/PD-L1 treatment **(B)** Kaplan Meier plot depicting overall survival of SUNY patients that received first-line anti-PD1/PD-L1 therapy stratified by PD-L1 IHC expression **(C-E)** Kaplan Meier plots depicting overall survival of first-line patients in PD-L1 negative (TPS < 1%), PD-L1 low (TPS = 1-49%) and PD-L1 high (>= 50%), cases. Significance of survival differences between distinct subgroups of patients was determined by the log-rank test.

When utilizing the TME status to predict treatment responses (i.e., predict immune inflamed as responder and immune desert as non-responder), we achieve a sensitivity of 0.46[95% CI: 0.35-0.58], specificity of 0.72[95% CI: 0.60-0.81] and positive predictive value of 0.64[95% CI: 0.51-0.76] for first line ICI-treated patients and a sensitivity of 0.44[95% CI: 0.35-0.54], specificity of 0.73[95% CI: 0.63-0.81] and positive predictive value of 0.63[95% CI: 0.52-0.74], when considering all ICI-treated patients. When utilizing PD-L1 expression to predict treatment response (i.e., predict PD-L1 >=50% as responder and PD-L1 1-49% or < 1% as non-responder), we achieve a sensitivity of 0.49[95% CI: 0.38-0.60], specificity of 0.74[95% CI: 0.63-0.84] and positive predictive value of 0.67[95% CI: 0.54-0.79] for first line ICI-treated patients and sensitivity of 0.44[95% CI: 0.35-0.54], specificity of 0.72[95% CI: 0.62-0.80] and positive predictive value of 0.63[95% CI: 0.51-0.73] when considering all ICI-treated patients. When combining TME status and PD-L1 expression into a single predictor, which defines any patient with either an inflamed TME or PD-L1 >= 50% as responders, we achieve a sensitivity of 0.69[95% CI: 0.58-0.79], specificity of 0.59[0.47-0.70] and positive predictive value of 0.64[95% CI: 0.54-0.74] for first line ICI-treated patients and a sensitivity of 0.65[95% CI: 0.56-0.74], specificity of 0.56[95% CI: 0.47-0.66] and positive predictive value of 0.62[95% CI: 0.52-0.70] when considering all ICI-treated patients. Collectively, these results highlight the complementary value of HistoTME for prognostication of NSCLC patients, especially for those with PD-L1 < 50% and being considered for first line treatment with ICI+chemotherapy.

### Interactions between TME signatures improve prediction of immunotherapy response

Interactions among the various components of the TME play a key role in influencing immunotherapy responses^38^. A previous study by Liu et al highlighted an example of this complexity, revealing in NSCLC patients treated with ICI that only high PD-L1 expression in macrophages was correlated with better overall survival, while high PD-L1 expression in tumor or stromal cells was not^39^. Hence, we developed a supervised ML model that incorporates interactions between H&E-inferred TME signatures to predict immunotherapy responders (See Methods).

Specifically, we engineered 1740 interaction features by taking the sum, difference, product, and quotient of each pair of signatures to characterize interactions between TME signatures, then used a random forest model to select the most important interaction features from the training set, and trained XGBoost, a gradient boosted decision tree, using selected interaction features for ICI response prediction **(Figure 6A**; Methods). After applying 5-fold cross-validation to the training set to optimize the number of features selected, 18 TME signature interactions were chosen. These interactions maximize the cross-validation receiver operating characteristic curve (AUROC), achieving a CV AUROC of 0.68 **(Supplementary Figure 11)**. Of the 18 pairwise interactions, coactivation molecules, T cell traffic, and MDSC traffic were incorporated most frequently **(Figure 6B).** We then retrained XGBoost on the full training set using these 18 selected interaction features. The trained model utilizing interaction features predicted response on the test set with an AUROC of 0.68 [95% CI: 0.55-0.80], whereas using TME signatures alone, without interactions, achieved an AUROC of 0.55 [95% CI: 0.41-0.69], although the difference was not significant (p=0.17, paired DeLong’s test) **(Figure 6C)**. Furthermore, when only considering patients from the test set that received first line ICI-treatment, we found accuracies for predicting treatment response improved to an AUROC of 0.75 [95% CI: 0.61-0.88], while achieving a lower AUROC of 0.51 [95% CI: 0.23-0.78] for non-first-line ICI-treated patients.

**Figure 6:**
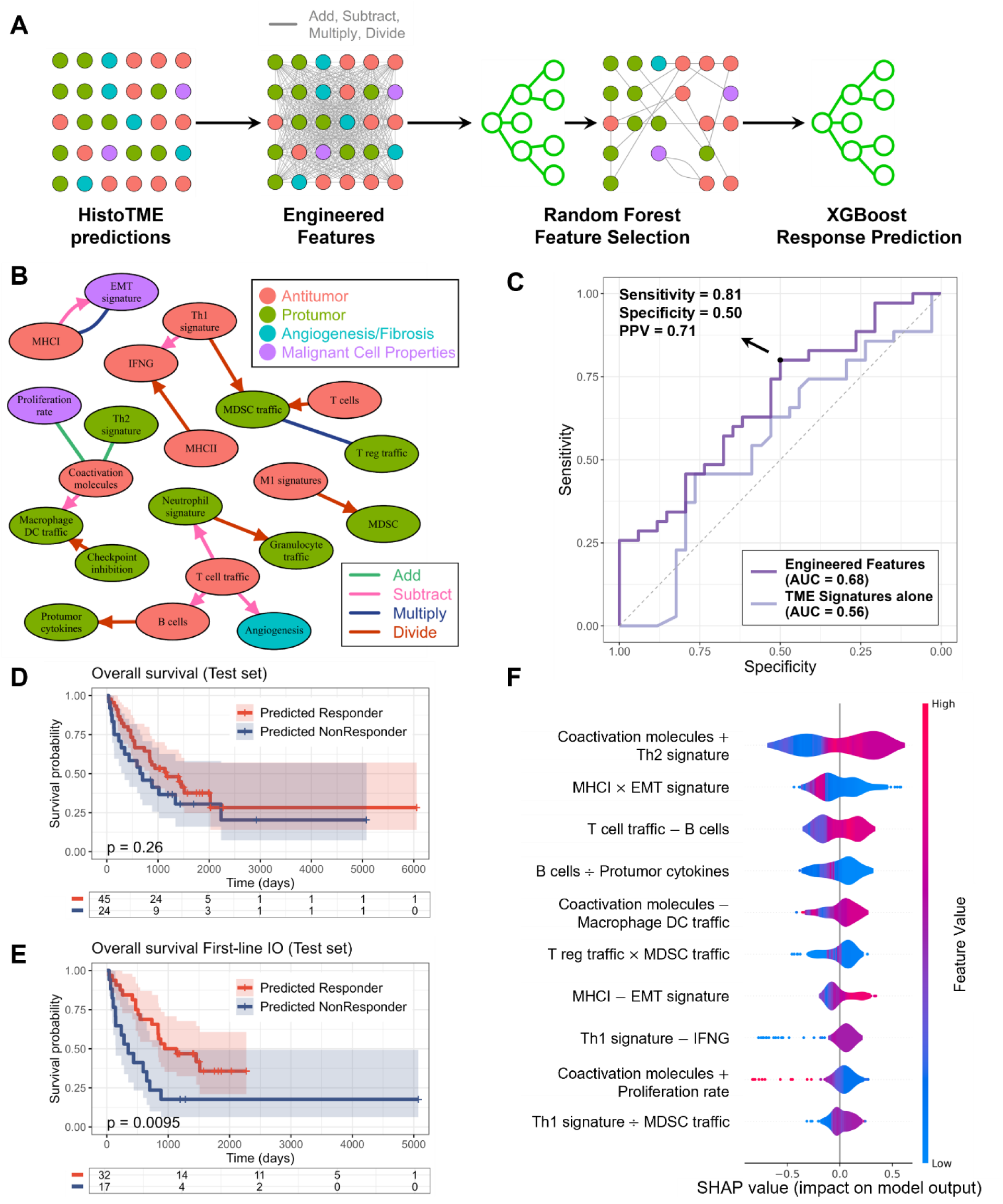
**(A)** Model development for response prediction: 1) HistoTME predictions are engineered into new features by taking pairwise sums, differences, products, and quotients. 2) random forest feature selection. 3) XGBoost trained for response prediction. **(B)** Feature network pairwise interactions of 18 selected features. Arrow endpoints denote the signature subtracted or divided from the signature at the start point. **(C)** Test set receiver operating characteristic (ROC) curve of the model trained on engineered features or TME signatures alone. Optimal cut point shown based on the Youden index. Kaplan Meier plot depicting overall survival of the test set stratified by AI response prediction for **(D)** all patients that received anti-PD1/PD-L1 treatment and **(E)** first-line immunotherapy (IO)-treated patients. **(F)** Shapley additive explanation (SHAP) summary plot ordered by SHAP importance.

At a probability threshold that maximized the Youden index, the model predicted responders with a sensitivity of 0.80 [95% CI: 0.63-0.92], specificity of 0.50 [95% CI: 0.32-0.68], and positive predictive value of 0.71 [95% CI: 0.49-0.87]. At this threshold, the predicted responders did not have significantly higher overall survival compared to non-responders in the entire test set (OS: HR = 0.35; 95% CI: -0.26 to 0.96; p = 0.26). However, when considering only first-line ICI-treated patients in the test set, the predicted responders had significantly longer survival time compared to non-responders (OS: HR = 0.90; 95% CI: 0.20 to 1.60; p = 0.0095) **(Figure 6D**, **6E)**. Furthermore, we observed significantly higher progression-free survival (PFS) outcomes for patients that were treated with first line ICI and predicted to be responders (PFS: HR = 0.94; 95% CI: 0.29 to 1.59; p = 0.0035) but not for second-line or subsequent-line ICI-treated patients **(Supplementary Figure 12)**. Using a Shapley additive explanation (SHAP)^40^ summary plot ordered based on feature importance, we determined TME signature interactions that had the most influence on the response prediction, with the most important interaction consisting of the sum between coactivation molecules and Th2 signatures (CM+Th2) **(Figure 6F)**. High levels of CM+Th2 corresponded to higher SHAP values, indicating higher predicted probabilities of response.

## Discussion

Immune checkpoint inhibitors have emerged as a promising treatment option for lung cancer. Yet only a minority of patients respond to these treatments, sometimes at the cost of severe toxicities and financial implications for those who may not respond to this therapy. Hence it is imperative to identify effective biomarkers capable of differentiating responders from non-responders prior to therapy initiation. The tumor microenvironment plays a fundamental role in shaping the responses of patients to immunotherapy. However, its dynamic nature, with multiple interacting components, makes it challenging to identify robust predictive biomarkers^32,33^. The integration of deep learning methods with digital pathology presents a promising and potentially cost-effective approach to interrogate the TME and alleviate some of these issues^41^.

In this work, we introduce HistoTME, a novel weekly supervised deep learning method to characterize the TME of patients from H&E slides, leveraging enhanced feature extraction capabilities of recent digital pathology foundation models. In contrast to recent deep learning approaches, which aim to predict spatially resolved gene expression profiles from histopathology images^42–45^, our approach aims to infer the TME composition through estimating the expression of distinct functional TME signatures. A key advantage of this approach is that by learning to predict the expression of gene signatures, we not only avoid overfitting to the expression of individual genes but also increase interpretability by directly relating specific histopathological features to previously established biological concepts^32^. Due to the limited benchmarking of foundation models for continuous biomarker prediction tasks, we experimented with three popular foundation models as feature extractors. We found out that the UNI foundation model^25^, when paired with multi-task AB-MIL, achieved the best predictive performance for the various TME signature prediction tasks. This improved performance likely stems from the considerably large histopathology datasets used to pre-train the UNI foundation model, as well as the added regularization induced by multitask learning.

With the help of HistoTME, we next classify patients into two distinct TME subtypes: Immune-Inflamed and Immune-Desert, characterized by distinct expression of anti-tumor and protumor immune signatures. These observations were further corroborated by significant differences in cellular abundances of T cells, B cells and macrophages within tumor tissue, based on matched IHC data of patients. Moreover, the two distinct TME subtypes were significantly predictive of OS and PFS in the patients treated with first-line immune checkpoint inhibitors. While the strongest evidence of ICI benefit in NSCLC can be attributed to patients with PD-L1 >= 50%, ICI has also been approved for patients with PD-L1 >= 1%, with the caveat that the benefit may not be as robust as the former group^2^. Our results suggest that predicted immune-inflamed TME may be utilized to better predict, and thereby select NSCLC patients for ICI treatment when PD-L1 is less than 50%, especially when considering ICI treatment in combination with chemotherapy.

In addition to clustering, we developed a novel supervised model to predict ICI response, which analyzes interactions between distinct TME signatures. Our supervised model achieves an AUROC of 0.68 in the unseen test patient cohort, improving to an impressive AUROC of 0.75 when predicting responses to first-line immunotherapy, highlighting the significance of interactions among various TME components in shaping treatment responses. On performing SHAP analysis, we found that the total expression of coactivation molecules and the Th2 signature were the most predictive of ICI response. This observation likely stems from the importance of coactivation molecules, such as CD28, which is essential for T cell activation after interacting with antigen-presenting cells, in cancer immunotherapy response^46^. Although Th2 signatures have been associated with an immune suppressive TME, their role in responses to ICI treatment remains unclear^47^. The emphasis laid by the AI model on this interaction could be explained by coactivation molecules promoting production of Th2 cytokines^48^, indicating that high levels of coactivation molecules and Th2 may offer a favorable environment crucial for therapeutic response. It is important to note, however, that interpreting the feature importance of ML models requires caution due to potential variability in feature importance depending on the training set, particularly with limited-size datasets.

Compared to previous studies of predicting response to ICI from H&E slides, we demonstrated a unique ability to identify responders using both unsupervised and supervised ML techniques. Both these approaches can be valuable for deriving ICI response biomarkers depending on the availability of clinically annotated datasets. Hu et al developed a supervised deep learning model that used extracted hidden features from histopathology images to predict anti-PD-1 response in melanoma and lung cancer. However, a main caveat of their approach was the limited interpretability of their hidden features^49^. In addition, they reported modest accuracies (AUROC: 0.645[95% CI: 0.495-0.784]) in predicting ICI response for lung cancer patients, which they attribute to the dataset consisting of core-needle biopsy samples rather than surgery samples. In another recent study, Wang et al achieved an impressive performance by using hand-crafted features derived from spatial interactions of tumor cells and TILs^20^; however, their approach required a minimum of five large image patches due to the reliance on spatial interactions, which may not generalize well on needle biopsies. Most importantly, to our knowledge, this is the first study showing that predicting molecular features of TME is feasible from scanned H&E images. This makes HistoTME extremely versatile and useful for analysis of both surgical resection and needle biopsy data. In fact, HistoTME signatures are able to effectively predict responses of patients while also maintaining interpretability, despite limited availability of tumor tissue from core needle biopsies, which make up ∼85% of the SUNY cohort.

This work has some limitations that should be further considered. First, although a large clinical cohort of ICI-treated patients was studied, we lacked additional external validation datasets to further validate the prediction of responses to ICI. Second, this work was conducted in a retrospective manner. We plan to further validate HistoTME in additional external cohorts of patients treated with ICI, both retrospectively and prospectively. Third, the development of HistoTME was limited to patients with NSCLC, implying that further testing and development will be required to extend the approach to different cancer types. Fourth, only three TME signature predictions were validated using IHC due to the lack of other strictly comparable IHC markers for other signatures. Bagaev et al^32^, who previously analyzed these TME signatures directly from transcriptomics data identified 4 distinct TME subtypes, which besides capturing differences in the activity of immune signatures also capture notable differences in activity of stromal and angiogenesis-associated signatures. In contrast, our approach results in the identification of two subtypes (Immune-Inflamed and Immune-Desert). This difference could potentially be explained by lower predictive accuracies of HistoTME for stroma and angiogenesis-associated signatures compared to immune signatures. We plan to utilize more complex molecular profiling tools, such as spatial transcriptomics, to further train and validate the accuracy of HistoTME predictions for other signatures. Lastly, while we showed promising results for predicting responses to first-line ICI treatment, the predictive accuracy of HistoTME was limited for the patients who received immune checkpoint blockade as second and subsequent-line treatment. This finding can be attributed to the fact that a majority of patients in this cohort received ICI as first-line treatment. In addition, since the TME is dynamically changing throughout the treatment, reflecting either response to therapy or tumor progression^50^, the H&E WSIs may not accurately represent the TME of patients following first-line treatment. Hence, it is important to consider the time interval between the H&E biopsy and ICI treatment to assess the utility of HistoTME for the prediction of treatment response.

In conclusion, HistoTME is an effective approach to characterize the TME of NSCLC patients and identify patients who will benefit from ICI therapy. Being based on H&E slides alone, HistoTME allows for a broad characterization of the TME without the need for expensive molecular tests or additional tissue stains. Given the routine use and low cost of H&E slides in diagnostic pathology along with the increasing adoption of digital and computation pathology in clinical practices, HistoTME promises to improve clinical management of cancer patients undergoing immunotherapy. Future research should focus on validating HistoTME in diverse patient populations and exploring its applicability to other cancer types, potentially extending its benefits beyond lung cancer. Finally, HistoTME can help advance our understanding of the role of TME in the context of other cancer treatments, opening avenues for the discovery of novel biomarkers and accelerate the adoption of personalized immuno-oncology.

## Data Availability

The TCGA Whole slide Imaging and bulk transcriptomics data is publicly available and can be downloaded from the GDC portal (https://portal.gdc.cancer.gov). The CPTAC lung cancer whole slide imaging data is publicly available at The Cancer Imaging Archive (https://www.cancerimagingarchive.net/collection/cptac-luad, https://www.cancerimagingarchive.net/collection/cptac-lscc), whereas the CPTAC lung cancer bulk transcriptomics data is available from the GDC portal. Whole slide imaging data from the SUNY Upstate cohort is currently not publicly available owing to restrictions of patient privacy. However, the data will be made available upon reasonable request. Full clinical metadata and HistoTME predictions for each cohort are available through Zenodo (https://zenodo.org/uploads/11490460).

https://wiki.cancerimagingarchive.net/display/Public/CPTAC+Imaging+Proteomics

## Acknowledgement and Funding Statement

This project was supported by an award from Upstate Foundation’s Hendricks Endowment. Data (digital images and clinical meta-data) from the institutional cohort was generated at the Pathology Research Core Lab using institutional resources and support.

## Code Availability

The codes to implement HistoTME are available at: https://github.com/spatkar94/HistoTME.git

## Methods

### Description of the SUNY NSCLC cohort

This retrospective institutional cohort was assembled based on the following criteria: primary diagnosis of non-small cell lung cancer who were followed at SUNY Upstate for at least a minimum of 2 years, and had PD-L1 testing record along with IHC slide availability including corresponding H&E slides (n=652 patients, 1329 H&E slides). The query and chart review were done using electronic medical records (EMR, EPIC) and pathology information system (Co-Path) for abstracting clinico-pathological, treatment and follow-up information. The study was reviewed by the ethics committee at SUNY Upstate and considered exempt from IRB oversight. The details of the cohort description are provided in **Supplementary Table 1.** Briefly, 292 (44.8%) patients were treated with immunotherapy, and of these patients, 230 had treatment response information (partial, complete, stable) as documented radiographically or clinically by the treating physician in the patient’s charts (progress notes, EPIC). More specifically, the responders were defined as patients that exhibited a partial, complete, or stable disease without experiencing any recurrence or death for at least 6 months since the start of ICI treatment. Non-responders were defined as patients who exhibited progressive disease or death within 6 months since the start of ICI treatment. Overall survival (OS) of patients was defined as the time from the date of diagnosis until death from any cause. Patients who were alive at the last follow-up were censored for overall survival analysis. Progression-free survival (PFS) of patients receiving checkpoint inhibitor treatment was defined as the time interval from immune checkpoint inhibitor start until progression or death. Patients who were alive without disease progression at their last follow-up were censored for PFS.

### Specimen availability, immunohistochemical analysis and slide scanning

Briefly, from 652 patients, 445 had tumor specimens available from primary disease sites (biopsy=398, resection=47) and 207 from metastatic sites (biopsy=169, resection=35). Briefly, serial immunostaining was done on four-micrometer-thick freshly cut serial sections using archived FFPE blocks of surgically resected specimens from 82 cases diagnosed with NSCLC (47 primary sites, 35 metastatic sites). The following biomarker panel was used for immune (CD3, CD20, CD4, CD8, CD163, FOXP3), cancer-specific (TTF-1, P40) and epithelial markers (Pac-CK). The staining was performed in a CLIA-certified clinical pathology lab using an automated immunostainer BenchMark Ultra (Roche Diagnostics, Germany) at SUNY Upstate Medical University. For pretreatment, antibody detection and counterstaining, the following reagents were used: ULTRA CC1 (Cat #950-124), UltraView DAB (Cat. 760-500), UltraView Red (Cat # 760-501) and Hematoxylin (Cat. 760-2021) according to the manufacturer’s instructions (Ventana Medical Systems; Roche Diagnostics, Germany). The details of the primary and secondary antibodies, antigen retrieval conditions, as well as detection methods are listed in **Supplementary Table 2**. PD-L1-stained slides (clone 22C3 PharmDx, Dako), along with negative controls and corresponding H&E of 406 patients, were requested from LabCorp and the rest (n= 246 patients) were obtained from the local pathology archives at SUNY Upstate Medical University. Glass slides were digitized using an Aperio AT2 Dx scanner (Leica Biosystems, CA, USA) at 40x magnification at the Pathology Research Core at SUNY Upstate. PD-L1 manual scoring was performed by expert pathologists using an FDA-approved assay and scoring guidelines at LabCorp. Tumor proportion score (TPS) was calculated as the % of viable positive tumor cells/all tumor cells, where positivity was defined as partial and/or complete membrane staining at any intensity (>1%) in tumor cells. PD-L1 quantification was categorized into clinically relevant groups as approved by the FDA: <1% (absent), 1-49% (low), and ≥50% (high).

### Description of Pre-processing Steps for Whole Slide H&E images

All WSI in the experiments were first preprocessed to mask out the tumor tissue from background using RGB to HSV color transformation, median blurring and Otsu thresholding^51^. Following tissue segmentation, each WSI was split into image tiles of physical size 256µm x 256µm (i.e 512 x512 pixels at 20x magnification). Each whole slide image tile with an overlap >25% with the tumor tissue was stain normalized using the Macenco algorithm^52^ and scaled to have 0 mean pixel intensity and standard deviation of 1 prior to being fed as input to an open-source foundation model—CTransPath^26^, RetCCL^30^ or UNI^25^—which learns to extract informative histopathologic features from each tile. CTransPath model consists of a convolutional neural network (CNN) and a multi-scale Swin Transformer architecture as its backbone, RetCCL uses a CNN-based architecture, and UNI implements a vision transformer (ViT). All foundation models were pre-trained self-supervised learning; CTransPath and RetCCL were pretrained on ∼30,000 WSIs, while UNI was pretrained on ∼100,000 WSIs. Feature extraction from each pretrained foundation model results in a feature matrix of shape *(n x 768)* for CTransPath, *(n x 2048)* for RetCCL, and (*n x 1024)* for UNI per patient, where n represents the number of total number image tiles derived from WSI of each patient.

### Experimental Setup and Implementation Details of HistoTME

HistoTME was trained using matched patient-level bulk RNA sequencing and whole slide imaging data of 865 patients (955 WSIs) from the TCGA cohort and validated on patient-level bulk RNA sequencing and whole slide imaging data of 333 patients (1501 WSIs) from the CPTAC cohort. Pre-processed bulk RNA sequencing data (gene level TPM counts) from each patient were downloaded from NCI Genomic Data Commons and further analyzed using the bioinformatics pipeline previously published by Bagaev et al to calculate the average normalized expression of distinct functional gene sets^32^, referred to as *TME signatures*, which comprehensively capture TME composition and its various functional characteristics.

For each patient, histopathologic features were extracted from WSI tiles using one of the three foundation models described above. WSI tile-level features were then concatenated together into a single “bag-of-features” representation to facilitate weakly supervised regression, using the attention-based multiple instance learning (AB-MIL) method proposed by Ilse et al^33^. The AB-MIL model consists of a learnable attention module, which assigns a weight, commonly referred to as *attention*, to each tile, and a feature aggregation module, which calculates the weighted sum of features across all tiles. This results in a single patient-level representation, summarizing key histopathological characteristics of the TME. The output of the feature aggregation module is then fed to a multilayer perceptron (MLP) module, which learns to predict the expression of a specific TME signature (single task) or multiple functionally related TME signatures (multi-task) as established previously by Ayers et al^31^ and Bagaev et al^32^. The AB-MIL model was trained with the AdamW optimizer^53^ and Huber loss function with delta set to one, which mitigates overfitting of model predictions to outliers by balancing the mean squared error and mean absolute error together, defined as follows:

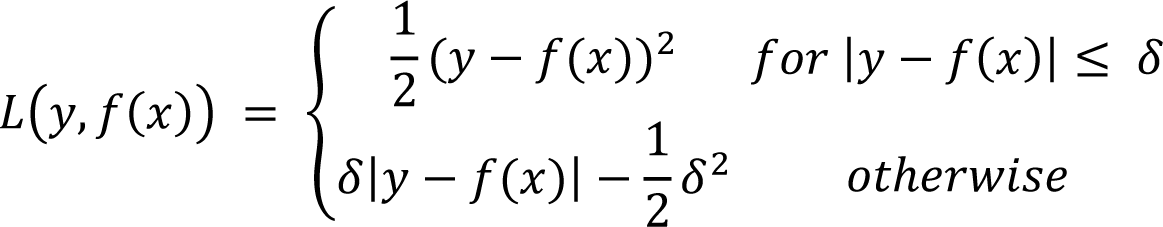

The learning rate was set to 1×10^-4^ with a weight decay of 1×10^-4^. Batch size was set to 1 with gradient accumulation for 8 batches. Overall, for each model benchmarked in this study, training was done for 40 epochs with early stopping criteria of 10 consecutive epochs with no improvement in validation loss.

After completion of training, the Pearson correlation metric was utilized to evaluate accuracy of predictions of each model on the independent validation set (CPTAC-NSCLC). 95% confidence intervals (CI) of the Pearson correlation metric were estimated through 1000 bootstrapping iterations using the SciPy package^54^. The model achieving the highest Pearson correlation coefficients, on average, was eventually selected for external testing and determining ICI efficacy. All AI models were developed using open source PyTorch version 2.1.0^55^.

The external test cohort consisted of serial H&E and multiplex IHC sections of surgically resected tumors from 82 NSCLC patients (47 primary tumors, 35 metastases) enrolled at SUNY Upstate Medical University (See cohort description above). The best model, as determined from the benchmarking experiments on the CPTAC validation set, was applied to this test cohort to estimate the expression of T cell, B cell and Macrophage signatures. These expression predictions were then compared against the actual cellular abundances of respective cell types from corresponding IHC slides, using both Pearson and Spearman correlation metrics given the different scales of quantification. Cellular abundances of T cells (CD4, CD8), B cells (CD20) and Macrophages (CD163) were estimated from corresponding IHC stains using an open-source cell counting software QuPath (v0.5.0)^56^. Specifically, for each whole-slide IHC image, the tumor tissue was manually segmented and separated from the background. Following manual segmentation, a standard pipeline of stain deconvolution and positive cell segmentation was implemented to quantify the cell type abundance, defined as the total number of marker-positive cells per mm^2^ (https://qupath.readthedocs.io/en/stable/docs/tutorials/cell_detection.html). Three patients from this test cohort contained surgical resections from lymph-node metastases with an extremely high number of immune cells and were excluded from correlation calculations, to avoid reporting inflated accuracies.

### Predicting TME status using HistoTME

K means clustering algorithm was utilized to cluster patients from the TCGA and CPTAC-NSCLC cohorts into distinct TME subtypes based on HistoTME-predicted microenvironment signatures. Optimal number of clusters for K means clustering was determined using the average silhouette score metric (**Supplementary Figure 4**), which revealed 2 distinct TME subtypes. Finally, a random forest classification model was trained on the clustered data to enable classification of individual patients from the SUNY cohort into distinct TME subtypes. To mitigate issues arising from data distribution shifts across cohorts, all predicted signature expression values were further scaled by their respective dataset-specific means and standard deviations prior to the development of the TME subtype classification model.

### Prediction of ICI Response Utilizing HistoTME-derived microenvironment signatures

Training and test sets were derived from 230 NSCLC patients with matched whole slide imaging data and treatment response labels as described above for supervised machine learning analysis using a stratified random splitting strategy (70% train, 30% test) implemented in scikit-learn package^57^. A total of 161 patients (84 responders, 77 non responders) were assigned to be part of the *IO training set* and 69 patients (35 responders, 34 non responders) were assigned to be part of the *IO test set*. A supervised machine learning model was trained to predict immunotherapy response using data from the IO training set. The model takes as input the 30 HistoTME predicted microenvironment signatures. In addition, 1740 interaction features were engineered by taking the sum, difference, product, and quotient of each pair of TME signatures. Since HistoTME signatures include negative and positive values, we take the exponent of each signature prior to computing the product and quotient to maintain monotonic relationships and preserve interpretability. A Random forest feature selection approach was used to select the top k most important features predictive of ICI response from the training set. A gradient-boosted decision tree model, XGBoost^58^, was trained using these selected features to predict ICI response. To identify the best set features for ICI response prediction, we utilized a 5-fold cross-validation strategy on the training data. Here, 80% of the training set was randomly allocated for model training, while the remaining 20% of the training set was reserved for validation purposes. A separate cross-validation iteration was conducted for each set of k features, and the number of boosting rounds was selected to optimize two metrics (minimize logistic loss and maximize AUROC) during cross-validation. Early stopping was set to 100 to select boosting rounds, where additional boosting rounds are not created if the metric does not improve after 100 rounds. The set of features that maximized the cross-validation AUROC was chosen as the final set of features. These features were then utilized along with XGBoost to train the final model on the entire training set. XGBoost learning rate was selected at 0.1, gamma was set to 0.1 to reduce overfitting, and the number of boosting rounds was set to 35 based on cross-validation. SHAP (Shapley Additive exPlanations) was used to interpret the output of the XGBoost model and estimate contribution of each feature^40^.

### Additional Analyses and Statistical Tests

The Wilcoxon ranked sum test was used to determine the significance of differences in cellular abundances, as inferred from IHC, between the two predicted TME subtypes. The log-rank test was used to evaluate the prognostic significance of HistoTME in predicting PFS and OS. Response prediction was evaluated by AUROC. 95% Confidence intervals for AUROC were computed by the DeLong approach^59^. The optimal cutpoint of the ROC curve was chosen at the threshold that maximized the Youden index (J)^60^. 95% confidence intervals of precision, recall, and positive predicted value of the response predictions were calculated using exact binomial confidence limits. Kaplan-Meier survival curves were used to visualize the differences in the OS and PFS of AI-predicted responders and non-responders. Hazard ratios between survival groups with 95% CIs were calculated using a univariate Cox proportional hazards regression model. All statistical tests were two-sided, with a p-value less than 0.05 considered statistically significant. All statistical analyses were performed in R version 4.3.1 unless otherwise specified.

## Supplementary Tables

**Supplementary Table 1** Summary of the clinical demographics of the SUNY Upstate NSCLC cohort. A total of 652 patients with whole slide imaging and/or clinical follow-up data were analyzed in this study. PD-L1 manual scoring was performed by expert pathologists using an FDA-approved assay and scoring guidelines at LabCorp. In an event where multiple biopsies were taken from a single patient, the overall PD-L1 score assigned to that patient was maximum of PD-L1 scores assigned to each biopsy. The full clinical table is accessible from the GitHub repository.

**Supplementary Table 2** Overview of reagents used for serial immunohistochemical staining of surgical resection specimens from the SUNY Upstate NSCLC cohort.

## Supplementary Figures

**Supplementary Figure 1:**
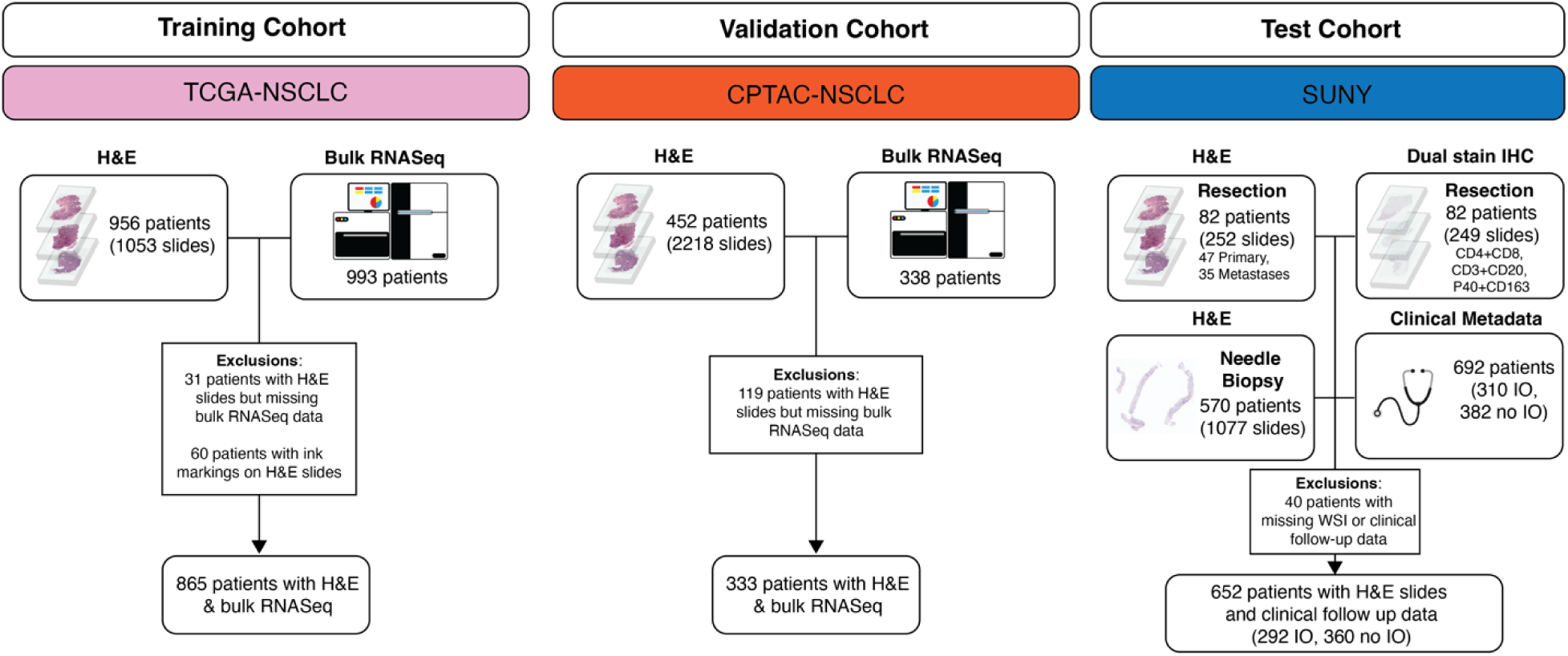
Overview of the study design. To train HistoTME matched whole slide imaging and bulk RNA sequencing data from 865 patients from TCGA was utilized. To validate HistoTME matched whole slide imaging and bulk RNA sequencing data from 333 patients from CPTAC was utilized. For testing HistoTME predictions, matched whole slide H&E imaging and immunohistochemistry imaging data from surgical resection specimens of 82 patients from SUNY Upstate Medical University was utilized. For evaluating ICI efficacy surgical resection or needle biopsy specimens from 290 patients of the test cohort with matched clinical follow-up data following ICI treatment was utilized.

**Supplementary Figure 2:**
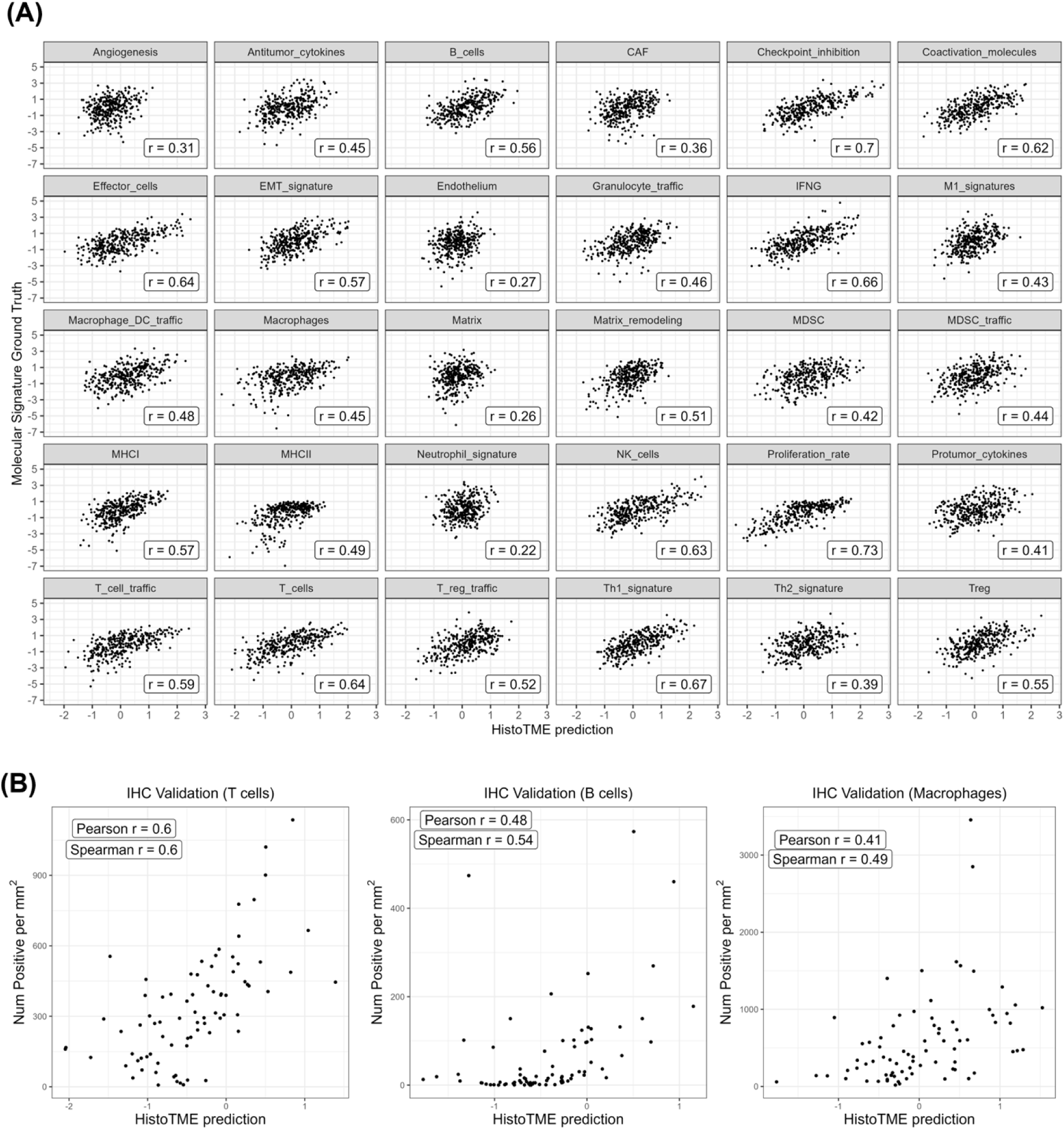
**(A)** Scatter plots between the observed values derived from transcriptomics and HistoTME predicted values is shown at the patient level on the CPTAC validation cohort (N=333). **(B)** Scatter plots between the cell type density, defined as the number of marker positive cells per mm^2^ from the immunohistochemistry (IHC) stain, and HistoTME predicted values is shown at the patient level on the external SUNY cohort (N=79). Cell type densities were quantified from whole slide immunohistochemistry images using QuPath v0.5.0 cell detection and classification algorithms and default parameters.

**Supplemental Figure 3:**
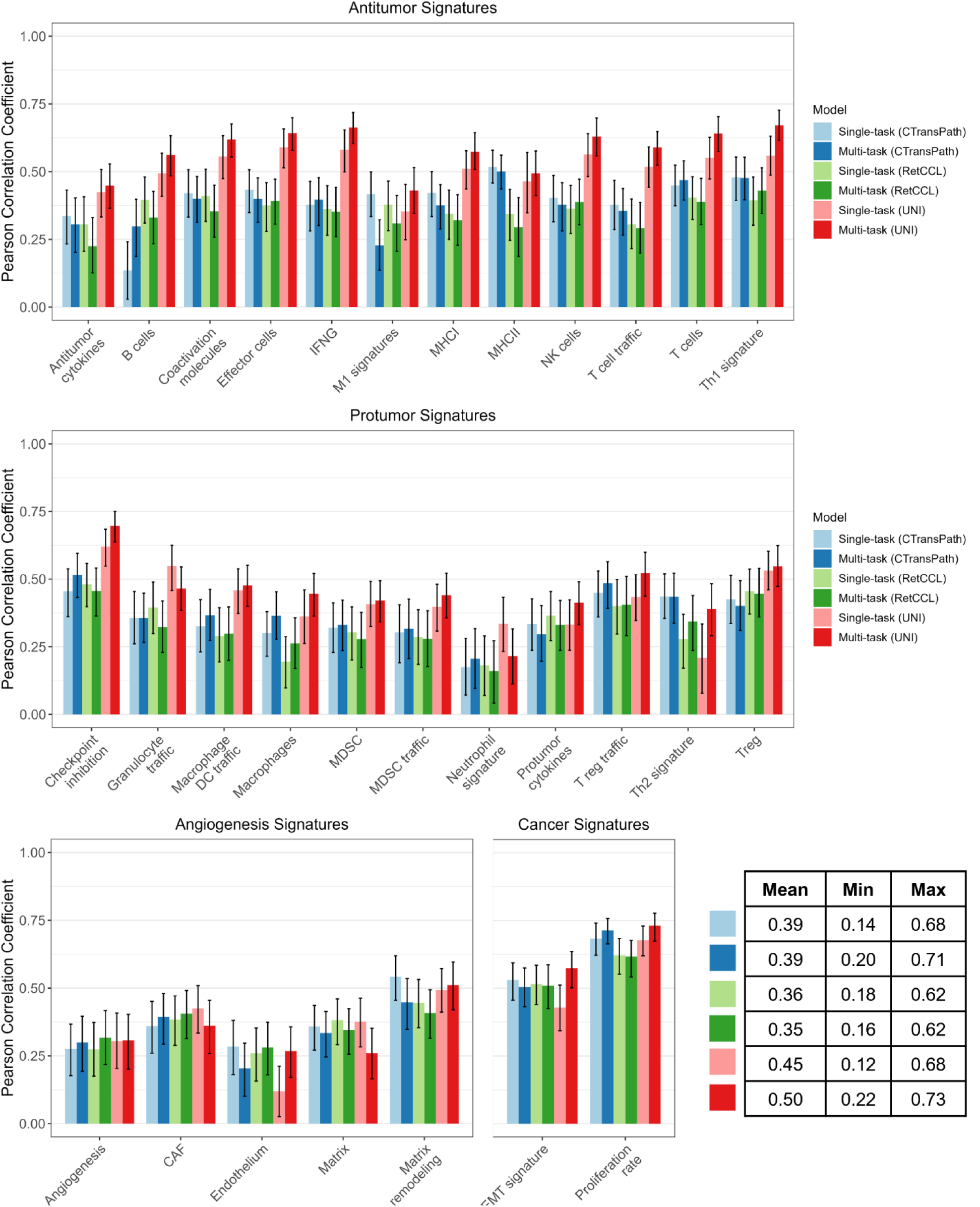
Pearson correlation between the observed values derived from transcriptomics and each model configuration’s predicted values is shown at the patient level on the external CPTAC validation cohort for antitumor, protumor, angiogenesis/stromal, and cancer/malignant cell-related signatures. Model configurations consisted of single-task and multi-task AB-MIL with CTransPath, RetCCL, or UNI as the feature extractor. Error bars represent the 95% confidence intervals. The mean, minimum, and maximum for each model across all TME signatures is shown in the table.

**Supplementary Figure 4:**
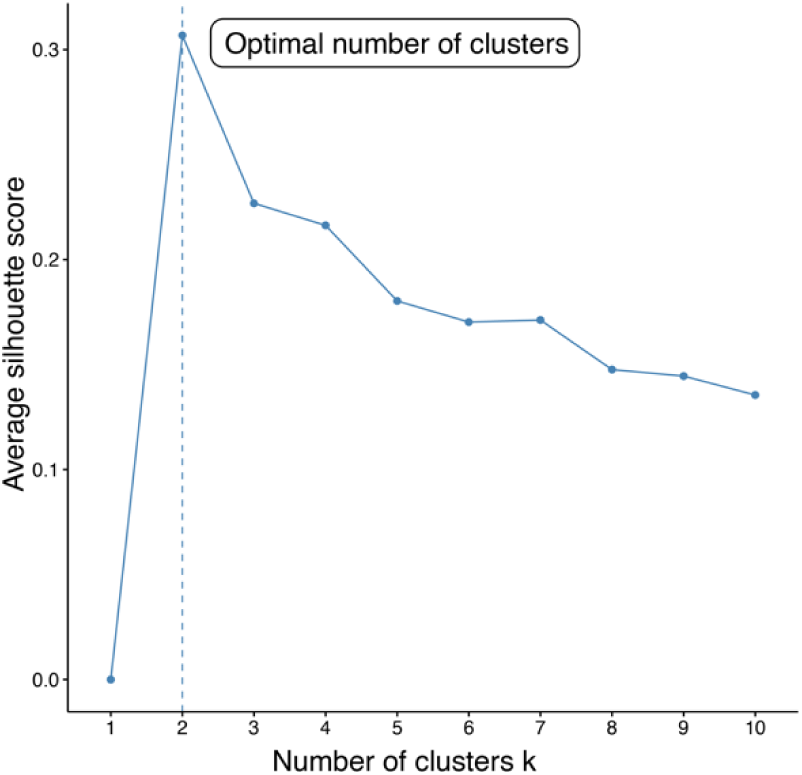
Silhouette score analysis to determine the optimal number of clusters from K means clustering. This analysis reveals K= 2 clusters maximize the average silhouette width. The input to the clustering algorithm were HistoTME-predicted expression of 30 TME signatures for TCGA+CPTAC-NSCLC patients

**Supplementary Figure 5:**
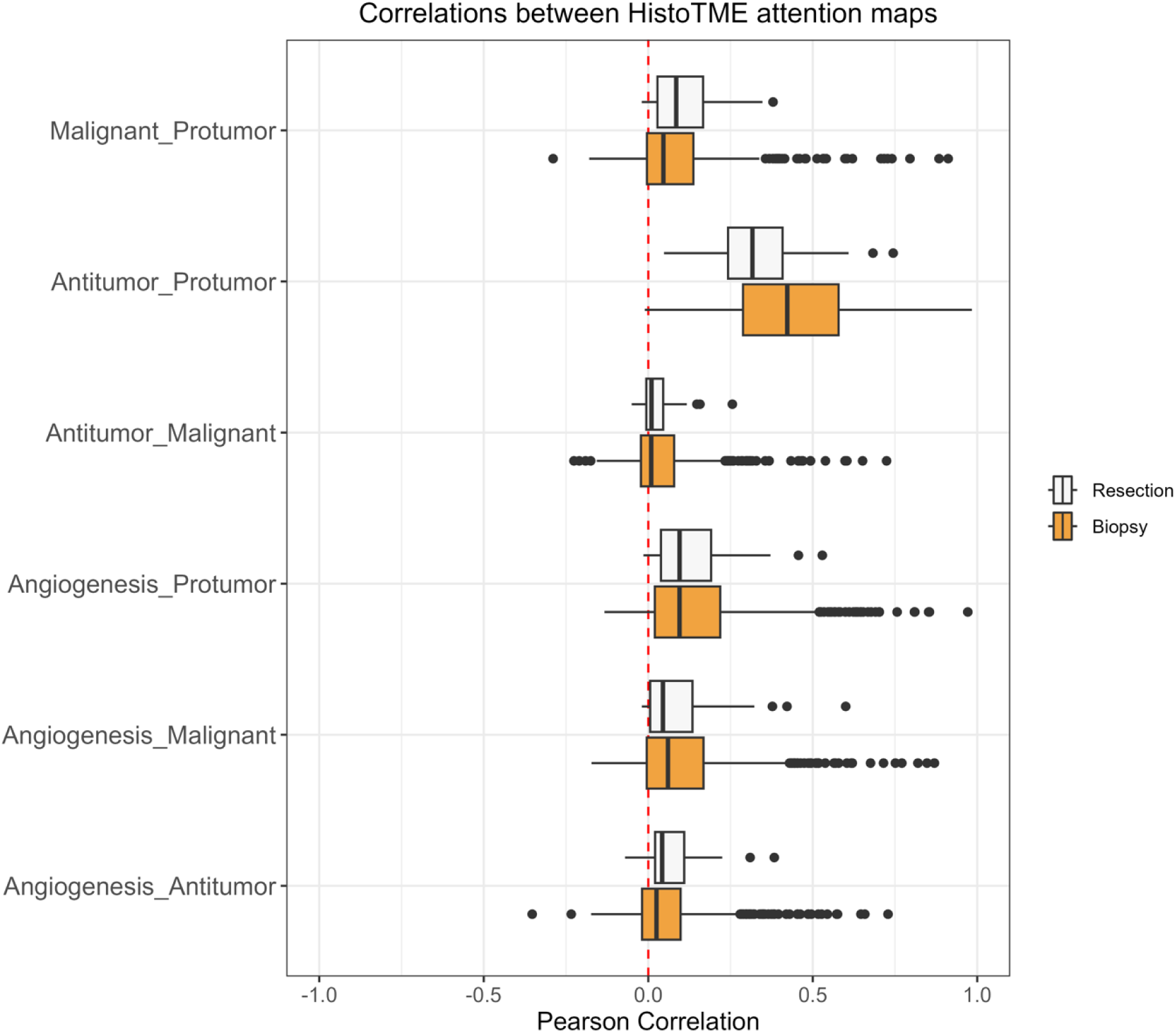
Distribution of pairwise Pearson correlations between attention maps for antitumor, protumor, angiogenesis/stroma, and malignant cell signatures, calculated on the SUNY cohort (652 patients, 1329 slides). Each data point in the distribution represents a single whole slide image.

**Supplementary Figure 6:**
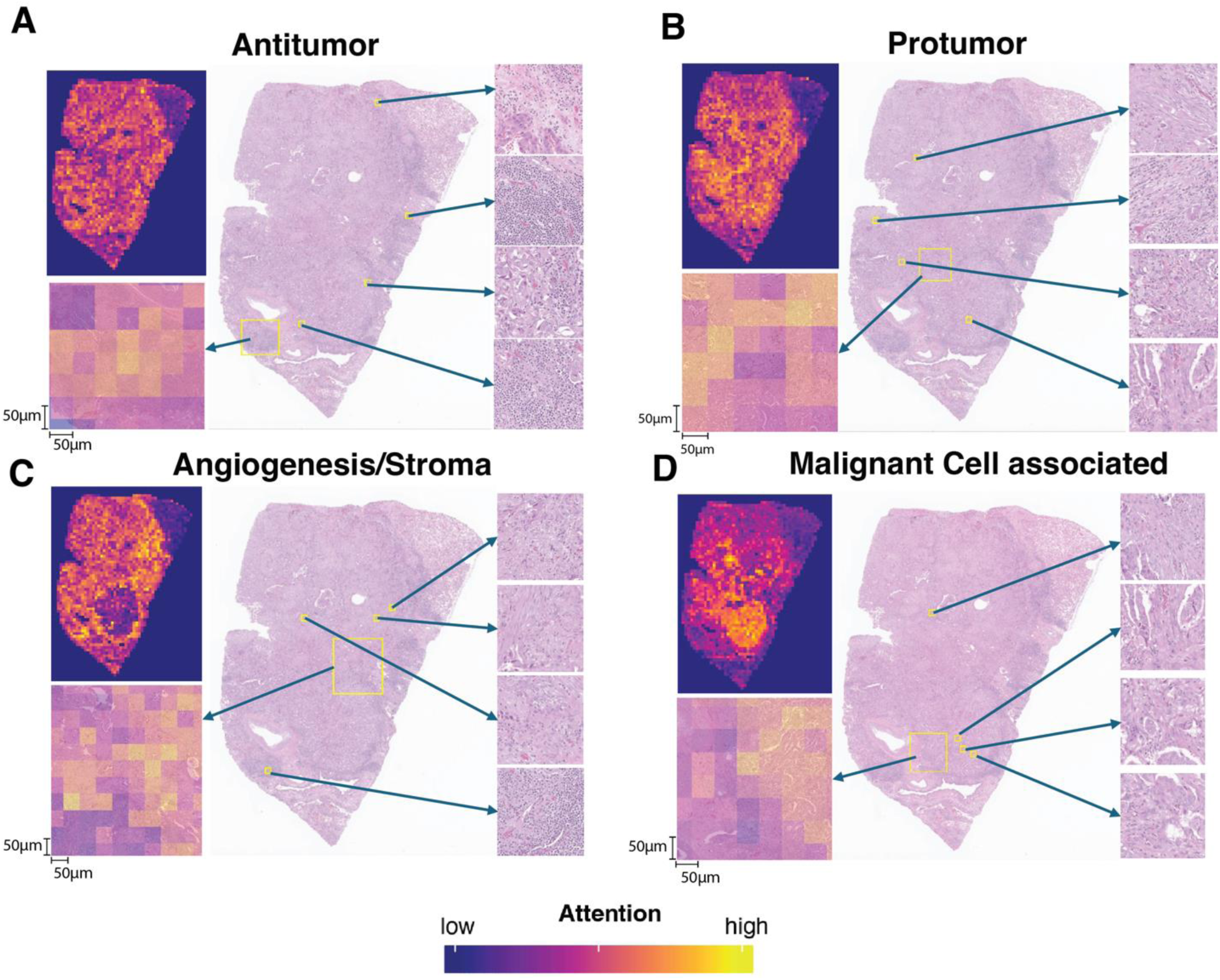
HistoTME-generated attention maps for a case predicted to have an Immune Inflamed TME. (A) attention maps highlighting regions of interest corresponding to antitumor immune signatures (B) attention maps highlighting regions of interest corresponding to protumor immune signatures. (C) attention maps highlighting regions of interest corresponding to angiogenesis/stroma-associated signatures (D) attention maps highlighting regions of interest corresponding to malignant cell-associated signatures.

**Supplementary Figure 7:**
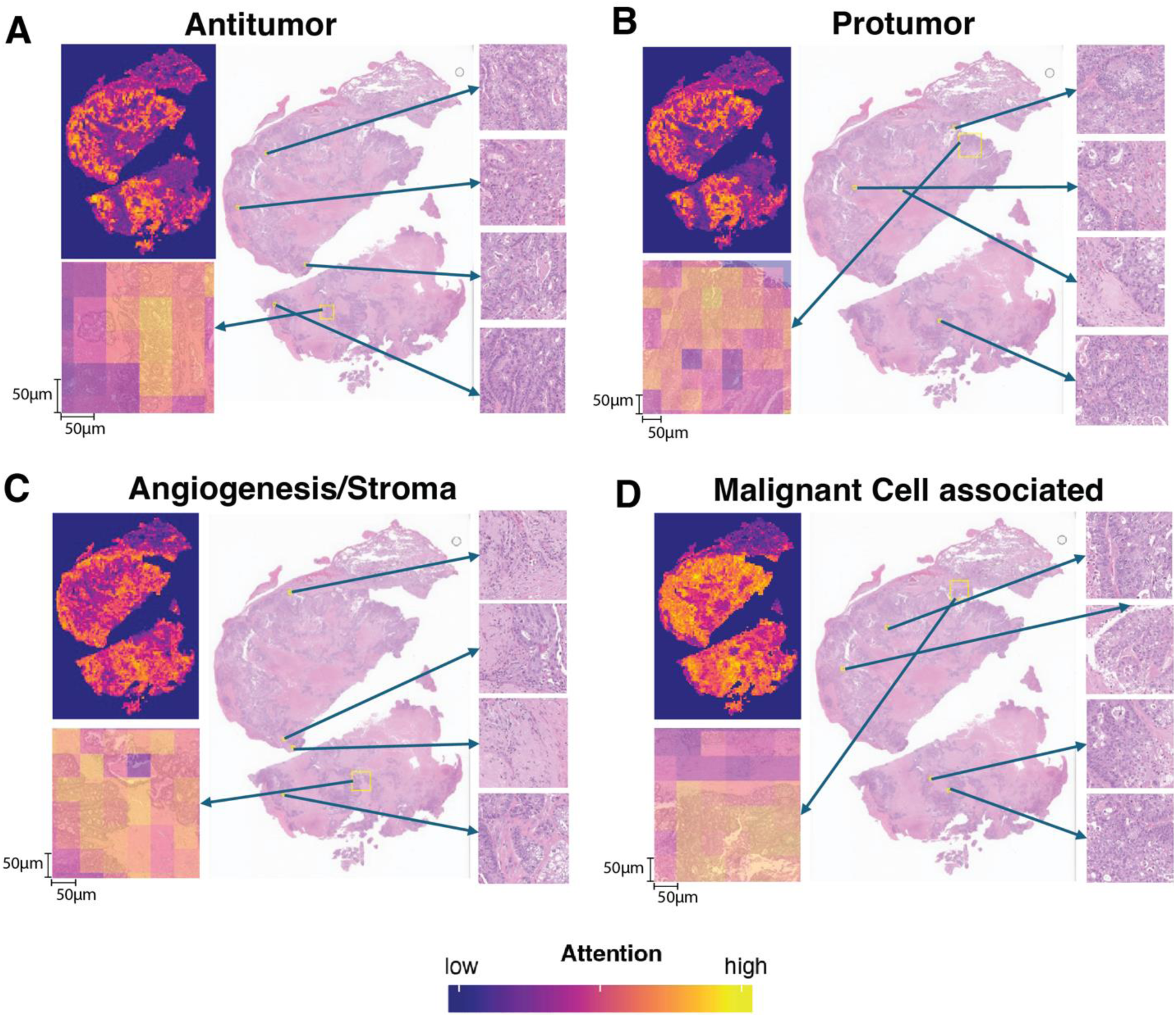
HistoTME-generated attention maps for a case predicted to have an Immune Desert TME. (A) attention maps highlighting regions of interest corresponding to antitumor immune signatures (B) attention maps highlighting regions of interest corresponding to protumor immune signatures. (C) attention maps highlighting regions of interest corresponding to angiogenesis/stroma-associated signatures (D) attention maps highlighting regions of interest corresponding to malignant cell-associated signatures.

**Supplementary Figure 8:**
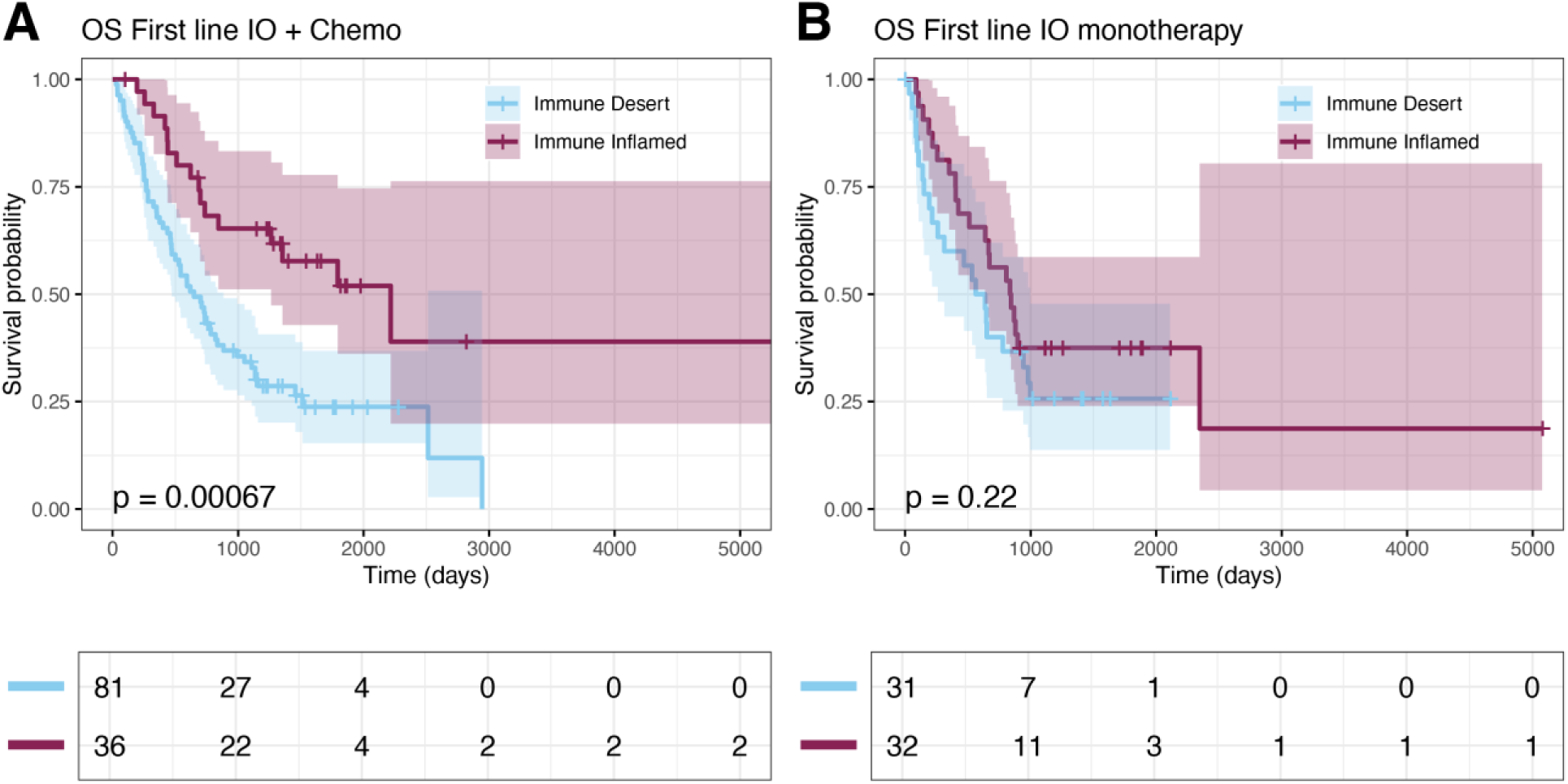
Association between HistoTME-based TME classification and overall survival outcomes of SUNY NSCLC patients treated with first-line ICI as combination therapy (first-line IO + Chemo) and monotherapy (IO monotherapy). **(A)** Kaplan Meier plot depicting overall survival−defined as time from date of diagnosis to date of death −of patients that received first-line IO + chemo **(B)** Kaplan Meier plot depicting overall survival of SUNY patients that received first-line IO monotherapy. Significance of survival differences between distinct subgroups of patients was determined by the log-rank test.

**Supplementary Figure 9:**
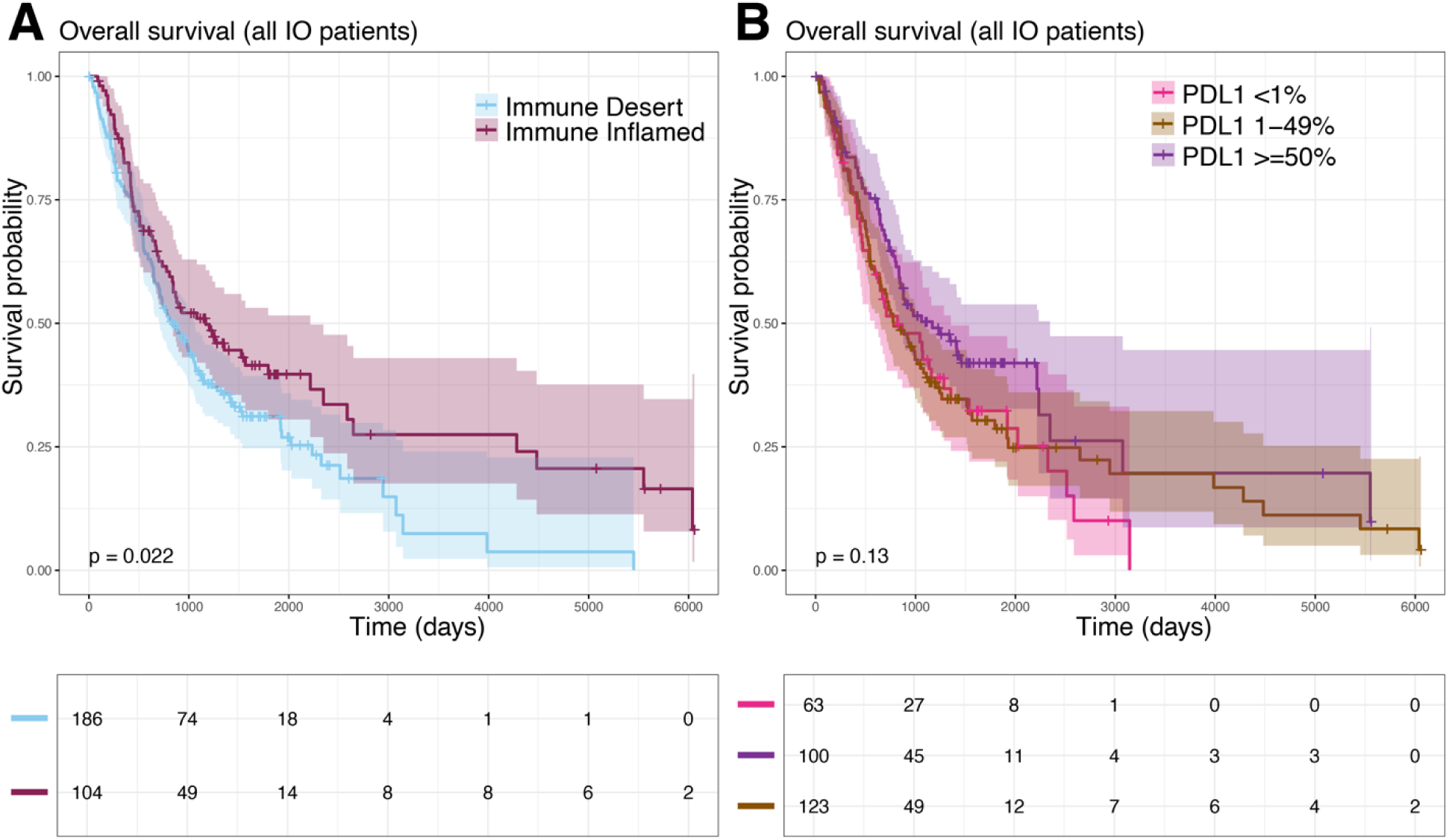
Overall survival (OS) of all immunotherapy (IO)-treated patients **(A)** stratified by Immune Inflamed and Immune Desert TME subtype **(B)** stratified by PD-L1 expression. Statistical significance between groups was estimated using the log-rank test.

**Supplementary Figure 10:**
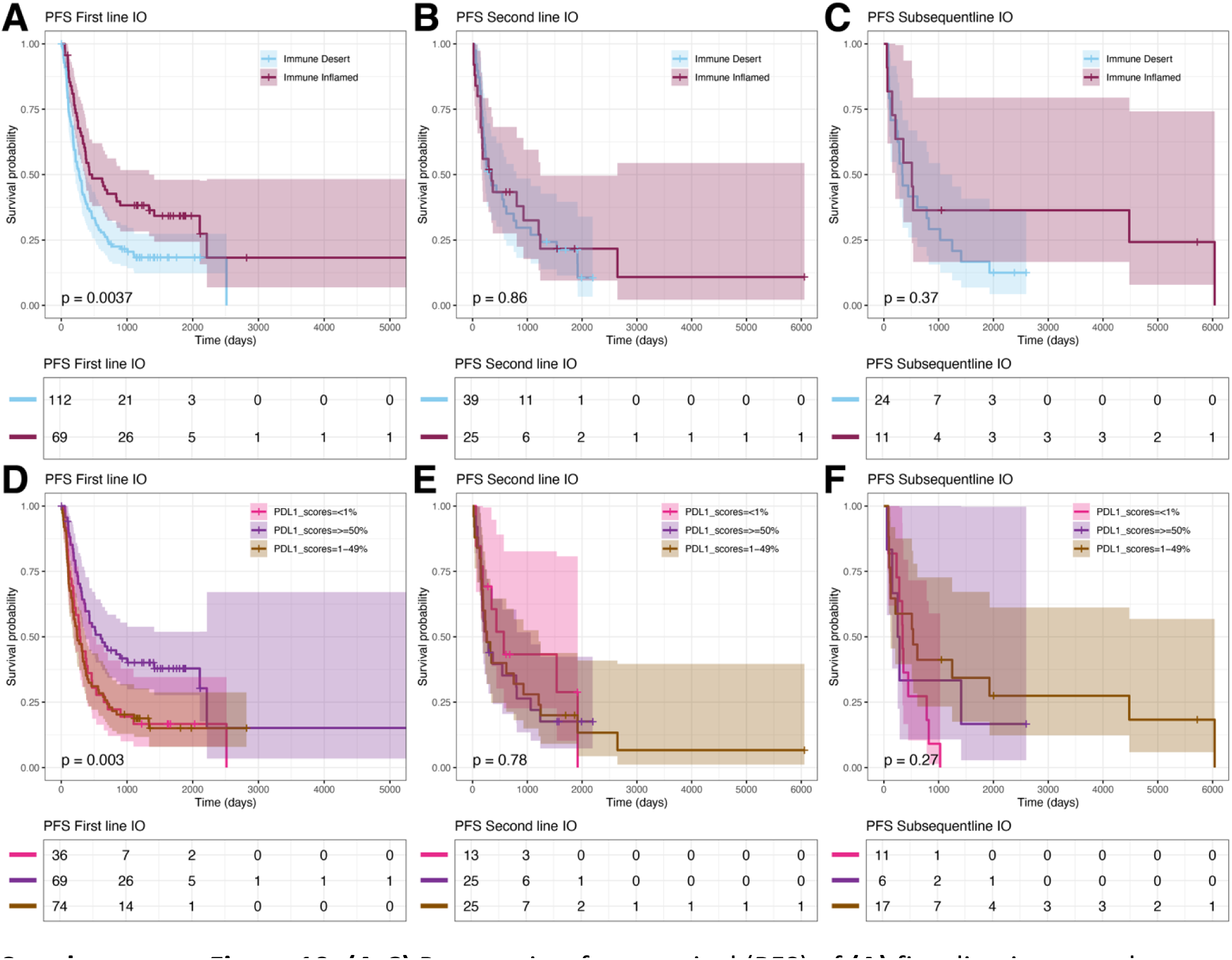
**(A-C)** Progression-free survival (PFS) of **(A)** first-line immunotherapy (IO)-treated, **(B)** second-line IO-treated, or **(C)** subsequent-line IO-treated patients in the SUNY Upstate cohort stratified by TME subtype. **(D-F)** Progression-free survival (PFS) of **(D)** first-line immunotherapy (IO)-treated, **(E)** second-line IO-treated, or **(F)** subsequent-line IO-treated patients in the SUNY Upstate cohort stratified by PD-L1 expression. Statistical significance between groups was estimated using the log-rank test.

**Supplementary Figure 11:**
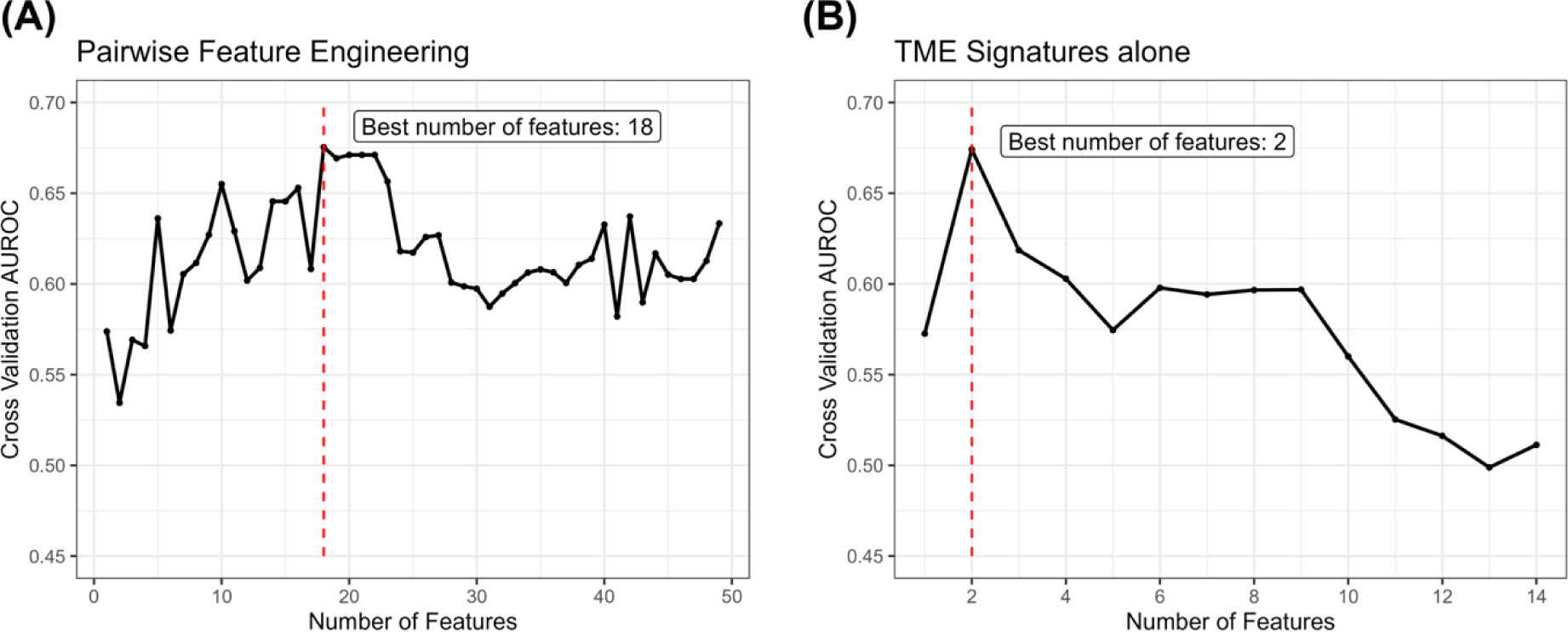
Random forest feature selection that maximized 5-fold cross-validation AUROC when XGBoost was trained to predict ICI response using (A) pairwise engineered TME signature interactions and (B) TME signatures alone. The number of features where AUROC was maximized was used.

**Supplementary Figure 12:**
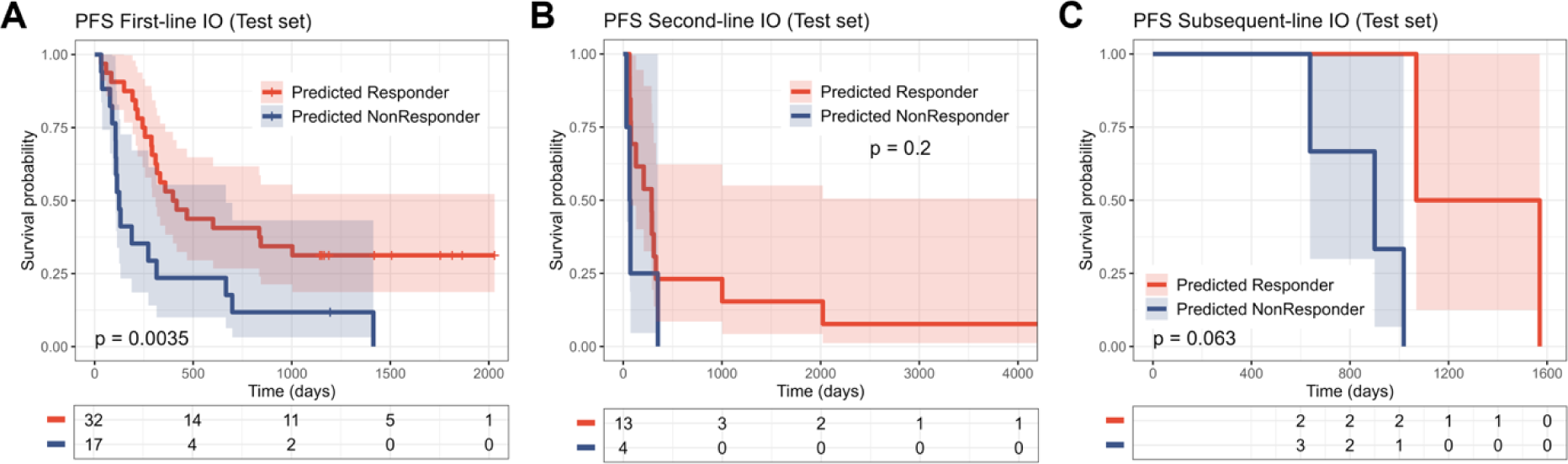
Progression-free survival (PFS) of **(A)** first-line immunotherapy (IO)-treated, **(B)** second-line IO-treated, or **(C)** subsequent-line IO-treated patients in the SUNY held-out test set (N=69) predicted to be responders or nonresponders. Statistical significance between groups was estimated using the log-rank test.

## References

1 Gridelli, C. et al. Non-small-cell lung cancer. Nat Rev Dis Primers 1, 15009 (2015). 10.1038/nrdp.2015.9

2 Sholl, L. M. et al. Programmed Death Ligand-1 and Tumor Mutation Burden Testing of Patients With Lung Cancer for Selection of Immune Checkpoint Inhibitor Therapies: Guideline From the College of American Pathologists, Association for Molecular Pathology, International Association for the Study of Lung Cancer, Pulmonary Pathology Society, and LUNGevity Foundation. Arch Pathol Lab Med (2024). 10.5858/arpa.2023-0536-CP

3 Rimm, D. L. et al. A Prospective, Multi-institutional, Pathologist-Based Assessment of 4 Immunohistochemistry Assays for PD-L1 Expression in Non-Small Cell Lung Cancer. JAMA Oncol 3, 1051–1058 (2017). 10.1001/jamaoncol.2017.0013

4 Chan, T. A. et al. Development of tumor mutation burden as an immunotherapy biomarker: utility for the oncology clinic. Ann Oncol 30, 44–56 (2019). 10.1093/annonc/mdy495

5 Hellmann, M. D. et al. Nivolumab plus Ipilimumab in Lung Cancer with a High Tumor Mutational Burden. N Engl J Med 378, 2093–2104 (2018). 10.1056/NEJMoa1801946

6 Marcus, L. et al. FDA Approval Summary: Pembrolizumab for the Treatment of Tumor Mutational Burden-High Solid Tumors. Clin Cancer Res 27, 4685–4689 (2021). 10.1158/1078-0432.CCR-21-0327

7 Jardim, D. L., Goodman, A., de Melo Gagliato, D. & Kurzrock, R. The Challenges of Tumor Mutational Burden as an Immunotherapy Biomarker. Cancer Cell 39, 154–173 (2021). 10.1016/j.ccell.2020.10.001

8 Tumeh, P. C. et al. PD-1 blockade induces responses by inhibiting adaptive immune resistance. Nature 515, 568–571 (2014). 10.1038/nature13954

9 Taube, J. M. et al. Colocalization of inflammatory response with B7-h1 expression in human melanocytic lesions supports an adaptive resistance mechanism of immune escape. Sci Transl Med 4, 127ra137 (2012). 10.1126/scitranslmed.3003689

10 Liu, Y. T. et al. Immune Cell PD-L1 Colocalizes with Macrophages and Is Associated with Outcome in PD-1 Pathway Blockade Therapy. Clin Cancer Res 26, 970–977 (2020). 10.1158/1078-0432.CCR-19-1040z

11 Yuan, Y. Spatial Heterogeneity in the Tumor Microenvironment. Cold Spring Harb Perspect Med 6 (2016). 10.1101/cshperspect.a026583

12 Lundberg, E. & Borner, G. H. H. Spatial proteomics: a powerful discovery tool for cell biology. Nat Rev Mol Cell Biol 20, 285–302 (2019). 10.1038/s41580-018-0094-y

13 Sorin, M. et al. Single-cell spatial landscapes of the lung tumour immune microenvironment. Nature 614, 548–554 (2023). 10.1038/s41586-022-05672-3

14 Cords, L. et al. Cancer-associated fibroblast phenotypes are associated with patient outcome in non-small cell lung cancer. Cancer Cell 42, 396–412 e395 (2024). 10.1016/j.ccell.2023.12.021

15 Mohanty, C. et al. SpatialView: an interactive web application for visualization of multiple samples in spatial transcriptomics experiments. Bioinformatics 40 (2024). 10.1093/bioinformatics/btae117

16 Ren, L. et al. Applications of single–cell omics and spatial transcriptomics technologies in gastric cancer (Review). Oncol Lett 27, 152 (2024). 10.3892/ol.2024.14285

17 Cao, L. et al. Deciphering spatial domains from spatially resolved transcriptomics with Siamese graph autoencoder. Gigascience 13 (2024). 10.1093/gigascience/giae003

18 Yu, Q., Jiang, M. & Wu, L. Spatial transcriptomics technology in cancer research. Front Oncol 12, 1019111 (2022). 10.3389/fonc.2022.1019111

19 Zhang, Q. et al. The spatial transcriptomic landscape of non-small cell lung cancer brain metastasis. Nat Commun 13, 5983 (2022). 10.1038/s41467-022-33365-y

20 Wang, X. et al. Spatial interplay patterns of cancer nuclei and tumor-infiltrating lymphocytes (TILs) predict clinical benefit for immune checkpoint inhibitors. Sci Adv 8, eabn3966 (2022). 10.1126/sciadv.abn3966

21 Saltz, J. et al. Spatial Organization and Molecular Correlation of Tumor-Infiltrating Lymphocytes Using Deep Learning on Pathology Images. Cell Rep 23, 181–193 e187 (2018). 10.1016/j.celrep.2018.03.086

22 Abousamra, S. et al. Deep Learning-Based Mapping of Tumor Infiltrating Lymphocytes in Whole Slide Images of 23 Types of Cancer. Front Oncol 11, 806603 (2021). 10.3389/fonc.2021.806603

23 Graham, S. et al. Hover-Net: Simultaneous segmentation and classification of nuclei in multi-tissue histology images. Med Image Anal 58, 101563 (2019). 10.1016/j.media.2019.101563

24 Diao, J. A. et al. Human-interpretable image features derived from densely mapped cancer pathology slides predict diverse molecular phenotypes. Nat Commun 12, 1613 (2021). 10.1038/s41467-021-21896-9

25 Chen, R. J. et al. Towards a general-purpose foundation model for computational pathology. Nat Med 30, 850–862 (2024). 10.1038/s41591-024-02857-3

26 Wang, X. et al. Transformer-based unsupervised contrastive learning for histopathological image classification. Med Image Anal 81, 102559 (2022). 10.1016/j.media.2022.102559

27 Lu, M. Y. et al. A visual-language foundation model for computational pathology. Nat Med 30, 863–874 (2024). 10.1038/s41591-024-02856-4

28 Azizi, S. et al. Robust and data-efficient generalization of self-supervised machine learning for diagnostic imaging. Nat Biomed Eng 7, 756–779 (2023). 10.1038/s41551-023-01049-7

29 Vorontsov, E., et al. Virchow: A million-slide digital pathology foundation model. *arXiv preprint arXiv:2309.07778* (2023).

30 Wang, X. et al. RetCCL: Clustering-guided contrastive learning for whole-slide image retrieval. Med Image Anal 83, 102645 (2023). 10.1016/j.media.2022.102645

31 Ayers, M. et al. IFN-gamma-related mRNA profile predicts clinical response to PD-1 blockade. J Clin Invest 127, 2930–2940 (2017). 10.1172/JCI91190

32 Bagaev, A. et al. Conserved pan-cancer microenvironment subtypes predict response to immunotherapy. Cancer Cell 39, 845–865 e847 (2021). 10.1016/j.ccell.2021.04.014

33 Ilse, M., Tomczak, J. & Welling, M. in International conference on machine learning. 2127–2136 (PMLR).

34 Pascal, L. E. et al. Correlation of mRNA and protein levels: cell type-specific gene expression of cluster designation antigens in the prostate. BMC Genomics 9, 246 (2008). 10.1186/1471-2164-9-246

35 Petitprez, F. et al. B cells are associated with survival and immunotherapy response in sarcoma. Nature 577, 556–560 (2020). 10.1038/s41586-019-1906-8

36 Chen, D. S. & Mellman, I. Elements of cancer immunity and the cancer-immune set point. Nature 541, 321–330 (2017). 10.1038/nature21349

37 Cristescu, R. et al. Pan-tumor genomic biomarkers for PD-1 checkpoint blockade-based immunotherapy. Science 362 (2018). 10.1126/science.aar3593

38 Sadeghi Rad, H., et al. Understanding the tumor microenvironment for effective immunotherapy. Med Res Rev 41, 1474–1498 (2021). 10.1002/med.21765

39 Liu, Y. et al. Immune cell PD-L1 colocalizes with macrophages and is associated with outcome in PD-1 pathway blockade therapy. Clinical cancer research 26, 970–977 (2020).

40 Lundberg, S. M. & Lee, S.-I. A unified approach to interpreting model predictions. Advances in neural information processing systems 30 (2017).

41 El Nahhas, O. S. M., et al. Regression-based Deep-Learning predicts molecular biomarkers from pathology slides. Nat Commun 15, 1253 (2024). 10.1038/s41467-024-45589-1

42 Schmauch, B. et al. A deep learning model to predict RNA-Seq expression of tumours from whole slide images. Nat Commun 11, 3877 (2020). 10.1038/s41467-020-17678-4

43 Hoang, D. T. et al. Prediction of cancer treatment response from histopathology images through imputed transcriptomics. Res Sq (2023). 10.21203/rs.3.rs-3193270/v1

44 He, B. et al. Integrating spatial gene expression and breast tumour morphology via deep learning. Nat Biomed Eng 4, 827–834 (2020). 10.1038/s41551-020-0578-x

45 Tan, X. et al. STimage:robust, confident and interpretable models for predicting gene markers from cancer histopathological images. bioRxiv, 2023.2005.2014.540710 (2023). 10.1101/2023.05.14.540710

46 Kamphorst, A. O. et al. Rescue of exhausted CD8 T cells by PD-1-targeted therapies is CD28-dependent. Science 355, 1423–1427 (2017). 10.1126/science.aaf0683

47 Jenkins, R. W., Barbie, D. A. & Flaherty, K. T. Mechanisms of resistance to immune checkpoint inhibitors. Br J Cancer 118, 9–16 (2018). 10.1038/bjc.2017.434

48 Skapenko, A., Lipsky, P. E., Kraetsch, H. G., Kalden, J. R. & Schulze-Koops, H. Antigen-independent Th2 cell differentiation by stimulation of CD28: regulation via IL-4 gene expression and mitogen-activated protein kinase activation. J Immunol 166, 4283–4292 (2001). 10.4049/jimmunol.166.7.4283

49 Hu, J. et al. Using deep learning to predict anti-PD-1 response in melanoma and lung cancer patients from histopathology images. Transl Oncol 14, 100921 (2021). 10.1016/j.tranon.2020.100921

50 Binnewies, M. et al. Understanding the tumor immune microenvironment (TIME) for effective therapy. Nat Med 24, 541–550 (2018). 10.1038/s41591-018-0014-x

51 Otsu, N. A Threshold Selection Method from Gray-Level Histograms. *IEEE Transactions on Systems*, Man, and Cybernetics 9, 62–66 (1979). 10.1109/TSMC.1979.4310076

52 Macenko, M. et al. in 2009 IEEE International Symposium on Biomedical Imaging: From Nano to Macro. 1107–1110.

53 Loshchilov, I. & Hutter, F. Decoupled weight decay regularization. *arXiv preprint arXiv:1711.05101* (2017).

54 Virtanen, P. et al. SciPy 1.0: fundamental algorithms for scientific computing in Python. Nature methods 17, 261–272 (2020).

55 Paszke, A. et al. Pytorch: An imperative style, high-performance deep learning library. Advances in neural information processing systems 32 (2019).

56 Bankhead, P. et al. QuPath: Open source software for digital pathology image analysis. Sci Rep 7, 16878 (2017). 10.1038/s41598-017-17204-5

57 Pedregosa, F. et al. Scikit-learn: Machine learning in Python. the Journal of machine Learning research 12, 2825–2830 (2011).

58 Chen, T. Q. & Guestrin, C. XGBoost: A Scalable Tree Boosting System. Kdd’16: Proceedings of the 22nd Acm Sigkdd International Conference on Knowledge Discovery and Data Mining, 785–794 (2016). 10.1145/2939672.2939785

59 DeLong, E. R., DeLong, D. M. & Clarke-Pearson, D. L. Comparing the areas under two or more correlated receiver operating characteristic curves: a nonparametric approach. Biometrics, 837–845 (1988).

60 Fluss, R., Faraggi, D. & Reiser, B. Estimation of the Youden Index and its associated cutoff point. Biom J 47, 458–472 (2005). 10.1002/bimj.200410135

